# The associations between recreational water contact, water quality measures, and acute gastrointestinal illness among Canadian beachgoers: a prospective cohort study

**DOI:** 10.64898/2026.04.01.26349959

**Authors:** Ian Young, Rachel Jardine, Binyam N. Desta, Thomas A. Edge, Faizan Saleem, David L. Pearl, Shannon E. Majowicz, Teresa Brooks, Andrea Nesbitt, J. Johanna Sanchez, Herb E. Schellhorn, Sarah Elton, Michael Schwandt, Dylan Lyng, Brandon Krupa, Elizabeth Montgomery, Mahesh Patel, Jordan Tustin

## Abstract

**Background:** Beaches are popular summertime destinations in Canada. However, they can be affected by specific fecal pollution sources, increasing the risk of recreational water illness.

**Objectives:** This study was conducted to determine the risks of acute gastrointestinal illness (AGI) among Canadian beachgoers and to evaluate the influence of different fecal indicator bacteria (FIB) and other water quality measures on assessing these risks.

**Methods:** In a prospective cohort design, beachgoers were recruited at sites across Canada from 2023 to 2025. Sociodemographic characteristics and exposures were determined through an on-site survey, with a 7-day follow-up survey to determine risks of AGI. Bayesian mixed-effects logistic regression models were fitted to evaluate the effects of an ordinal water contact variable (no contact, minimal contact, body immersion, and swallowed water) on the incident risk of AGI, with an interaction included for water quality indicators. The levels of six FIB and water quality measures were assessed: *Escherichia coli*, enterococci DNA, three microbial source tracking DNA markers (human HF183/BacR287, human mitochondria, seagull Gull4), and turbidity.

**Results:** A total of 4085 participants were recruited, with 67.6% completing the follow-up survey. The overall incident risk of AGI was 2.6%. Both swallowing water and body immersion increased AGI risks compared to no water contact: median of 20 excess cases (95% Credible Interval [CrI]: 4, 64) and 5 excess cases (95% CrI: 1, 19) of AGI predicted per 1000 beachgoers, respectively. *Escherichia coli* and seagull DNA marker levels were associated with AGI among those who had water contact, particularly among those who reported swallowing water.

**Discussion:** While the overall burden of AGI due to beach water contact in Canada was low, increased risks are associated with *E. coli* levels particularly among those who swallow water. This could be related to fecal contamination from seagulls. However, there is substantial uncertainty in the predicted effect sizes.

## 1. Introduction

Swimming and other recreational water activities are popular in Canada during the summer months. For example, a national survey conducted from 2023-2024 found that 25% and 21% of people living in Canada reported swimming in natural water bodies within the past seven days during the warmest months of July and August, respectively ^1^. These exposures are more common among children and youth (aged ≤19 years) compared to adults (∼10% vs. 6%) ^1^. Unfortunately, contamination of beach water from human sewage and animal feces can increase the risk of recreational water illness among beachgoers ^2–6^. In particular, exposure to fecal contamination in beach water increases the risk of acute gastrointestinal illness (AGI) ^2,4,7,8^. For example, a systematic review of global evidence found that swimming is associated with a 2.2 times (95% CrI: 1.82, 2.63) higher risk of AGI compared to beachgoers who do not have any water contact ^2^. Human sewage and wastewater effluent can contain specific enteric pathogens that are the primary etiological agents for AGI among beachgoers, particularly enteric viruses (e.g., norovirus, adenovirus) ^9–11^.

Children and youth are disproportionately affected by recreational water illness as they have developing immune systems, spend more time playing in the water, and are more likely to swallow water during water contact ^3,4,12–15^. Prior US cohort studies found that the risk of AGI due to swimming in beach water among children aged 0-4 years was more than double that of those aged 10+ years (22.4 vs. 7.8 episodes per 1,000 beachgoers) ^4^. While estimates are not available for the cost of recreational water illness in Canada, 90 million cases are estimated to occur each year in the US, resulting in an economic burden of US$2.2–3.7 billion annually ^8^.

In Canada, guidelines for recreational water quality are published by Health Canada, with the most recent guidance on fecal indicator bacteria (FIB) and beach water monitoring released in 2023 ^16,17^. These guidelines are available for adoption by local and provincial public health and environmental authorities when developing recommendations for recreational water use, such as the routine monitoring of beaches for FIB and informing beachgoers of health risks throughout the swimming season ^17^. The Beach Action Values (BAVs) in these national guidelines are based on US epidemiological studies because there is a lack of primary research in Canada on the burden of recreational water illness. Only one prior Canadian beach cohort study was conducted in Ontario in 1980 ^18^. Given that fecal contamination levels and risks of recreational water illness are affected by diverse pollution sources, environmental conditions and weather patterns, and beachgoer activities ^2,19–21^, there was a need to conduct a national prospective cohort study to determine risks of recreational water illness in Canadian beach settings. The aims of this study were to inform the national recreational water quality guidelines and local and provincial risk management decisions with Canada-specific data. The larger study, called the Canadian Beach Cohort Study, addressed several specific objectives as reported in a previously published protocol ^22^. This article reports on one of these objectives: estimating the risk of AGI among beachgoers by water contact exposure and levels of FIB contamination and other water quality measures.

## 2. Methods

### 2.1 Study design and settings

The study protocol was previously published and registered at ClinicalTrials.gov (ID: NCT06413485) ^22^. A prospective cohort design was used similar to prior US studies ^5,12,23^, and was piloted for feasibility in Toronto, Ontario, in 2022 ^14^. Briefly, the design included recruiting and surveying beachgoers at targeted beach locations, across four Canadian provinces, in British Columbia, Manitoba, Ontario (Toronto and Niagara Region), and Nova Scotia from 2023–2025. Specifically, the study was conducted at two beaches in Toronto in 2023, two beaches in each of Vancouver, British Columbia, and Grand Beach Provincial Park, Manitoba, in 2024, and two beaches in Niagara Region, Ontario, and Halifax, Nova Scotia, in 2025. Recruitment was completed over 33-36 days per site each summer between June and September. The beaches within each site were selected with regional partners based on their popularity with beachgoers and water quality history. All beaches were known to be affected by various (specific) non-point fecal pollution sources ^22^.

The members of each recruited household were surveyed to determine their characteristics and beach exposures. These surveys were completed on study tablets using a web-based platform (SimpleSurvey, OutSideSoft Solutions Inc., Quebec, Canada). Participants were asked to complete a follow-up survey seven days later by text message, email, or telephone to report any illness outcomes experienced after their beach visit.

### 2.2 Study population and eligibility

The study population included beachgoers of any age present at the beaches during recruitment days. Additionally, beachgoers had to live in Canada or the US at the time of recruitment, and they needed to be able to complete the survey in English or French. Participants living in the same household were recruited and surveyed together by trained research assistants. After completing a screening form, each eligible participant provided informed consent through a digital form, with parents and guardians aged 16 or older providing consent for any children and youth in the household. Given the acute and self-limiting nature of the health outcomes, individuals were able to participate again after a 21-day washout period ^6,12,23^.

### 2.3 Exposures and outcomes of interest

The primary exposure of interest was level of recreational water contact. An ordinal exposure variable was constructed with an expectation of identifying a dose-response relationship: 1) no water contact (unexposed group); 2) minimal contact; 3) body immersion; and 4) swallowed water ^3,4,24,25^. Minimal contact was defined as water contact without body immersion (e.g., wading in water below one’s waist), body immersion included entering the water above one’s waist (e.g., swimming), and swallowing water as ingesting any amount of water regardless of the level of body immersion. Participants were classified into one of these four categories based on their responses in the survey. The primary outcome of interest, and the focus of this report, was incident AGI self-reported within the 7-day follow-up period. AGI was defined as one or more of: (a) diarrhea (≥3 loose stools within 24 hrs); (b) vomiting; (c) nausea with stomach cramps; or d) nausea or stomach cramps that also caused missed work or school ^4–6,12,23^.

### 2.4 FIB and Water Quality Measures

At beach sites visited in 2024-2025, two water samples were collected from disctinct locations at each beach on each recruitment day at ∼12PM local time and analyzed for the following FIB and water quality measures: generic *E. coli* by standard culture ^26,27^, microbial source tracking (MST) DNA markers (human HF183/BacR287, human mitochondria, and seagull Gull4) using the ThermoFisher QuantStudio Absolute Q digital PCR system ^28,29^, enterococci using the U.S. EPA-1609.1 qPCR method ^30,31^, and water turbidity by nephelometry in the lab or handheld turbidimeter. Samples for molecular analysis were filtered (100 mL for enterococci qPCR and 300 mL for MST analysis) within 6-8 hours of collection by a local accredited lab in each province, and filters were then frozen and held at either −80°C (when possible) or −20°C for up to three months for molecular analysis. Samples for *E. coli* culture were filtered and incubated within 24 hours upon delivery to the local labs. A slightly different process was used in Toronto in 2023 due to the study only receiving preliminary funding at that time: *E. coli* data were obtained from the local public health unit using their collection time and approach (5-6 samples per beach collected at ∼7AM each day), samples for MST analysis were collected at ∼10AM and were filtered and analyzed within 24 hours (without freezing), and enterococci analyses were not conducted.

Some assumptions and data substitutions were needed for the *E. coli* data prior to analysis due to limit of detection issues. A lower limit of detection of <10 CFU/100 mL was used by labs in Toronto (7% of samples across 5/35 days) and Manitoba (20% of samples across 10/35 sample days), counts with this value were replaced with a mid-point value of 5 CFU/100 mL. Additionally, the Toronto lab had a maximum upper limit of reporting counts at 1000 CFU/100 mL which affected 11 samples on three recruitment days. These values were set at a value of 1001 CFU/100 mL as the most conservative estimate. Due to technical issues, two sample days from Manitoba consisted of only one sample for the molecular analysis.

### 2.5 Modelling framework and description

A series of multilevel logistic regression models were fitted to assess the research objectives. A Bayesian framework was used to incorporate uncertainty in all model parameters to allow direct probability-based inferences based on the observed data ^32–34^. The outcome for all models was incident AGI (yes or no). The unit of analysis was individual participants. The models evaluated the effect of level of water contact as the primary exposure of interest, including its interaction with levels of FIB and other water quality measures. The models were constructed using a Bernoulli distribution with hierarchical random intercept effects for location-recruitment date, beach, and site ^32^. The water contact variable contained four ordered categories of exposure, and therefore was modelled as a monotonic ordinal predictor ^35^. Briefly, this means that the model estimated simplex parameters representing differences between each ordered category (e.g., body immersion vs. minimal contact), as well as a beta parameter reflecting the average difference between adjacent categories ^35^. First a model containing only the water contact exposure and random intercept effects was fitted, then models containing the FIB-water quality measure interaction, and minimal sufficient adjustment set of confounding variables were fitted in an iterative process to arrive at the maximal model described below. The model equation, parameters, and prior distributions are shown below:

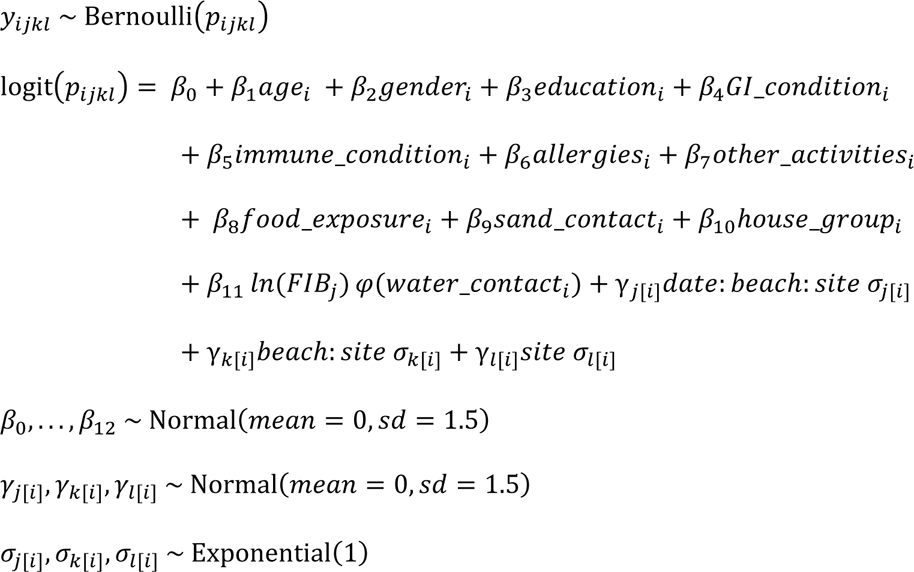

The outcome 𝑦_𝑖𝑗𝑘𝑙_ refers to the AGI status of each participant 𝑖 recruited on the 𝑗th location-recruitment day from the 𝑘th beach and 𝑙th site, and 𝑝_𝑖𝑗𝑘𝑙_ refers to the probability of a participant reporting AGI. The global intercept is represented by 𝛽_0_ and each confounding variable 𝛽_1_,…, 𝛽_10_ is represented by 𝛽 parameters, with categorical variables modelled as one or more separate indicator variables, each with one referent category. The 10 confounding variables included: age group (0-4 [referent], 5-9, 10-14, 15-19, 20+); gender (man/boy [referent], woman/girl, trans/gender fluid); highest education in household (high school or less [referent], college or trades, bachelor’s degree, post-graduate degree); presence of a chronic GI condition (yes, no [referent]); immune-compromised (yes, no [referent]); presence of allergies (yes, no [referent]); swam or engaged in other recreational water activities in the past 14 days (yes, no [referent]); consumed food at the beach (yes, no [referent]); played in the sand at the beach (yes, no [referent]); and participated in the study with at least one other household member (yes, no [referent]). These variables were identified through a directed acyclic graph and represent the minimal sufficient adjustment set of confounding variables for the water contact-AGI relationship (Figure S1); the graph was constructed based on reviewing prior beach cohort studies and discussion with the study team and partners ^22^.

The exposures of interest were represented by the log transformed 𝛽_11_𝐹𝐼𝐵 and the monotonic effect for level of water contact, represented by 𝜑. The monotonic effect simplex parameter was given a prior of Dirichlet(1, 2, 3), reflecting an expectation of a dose-response relationship ^35^. The random intercepts are represented by the 𝛾 parameters, with their variation represented by 𝜎. We specified weakly informative prior distributions (priors) for model parameters so that the model would be skeptical of highly implausible values ^32,33^. All 𝛽 parameters were given priors with a normal distribution (mean = 0, SD = 1.5). The 𝜎 parameters were given exponential priors with a rate of 1 ^32^.

The first constructed model (Model 1) included culture-based *E. coli* in CFU/100 mL as the FIB measure. Additional models were constructed and evaluated to compare the following different FIB and water quality measures: human marker (HF183/BacR287) as number of DNA copies/100 mL (Model 2); human mitochondrial DNA copies/100 mL (Model 3); seagull feces (Gull4) marker DNA copies/100 mL (Model 4); enterococci in calibrator cell equivalents/100 mL (Model 5); and water turbidity in NTUs (Model 6). Two different formulations for the FIB and water quality measure models were compared: a) average of the two samples per beach and b) highest single sample value at each beach location. As enterococci data were not available from Toronto, data from that site were excluded from the model evaluating this FIB measure (Model 5). Additionally, *E. coli* data from Toronto were summarized as a geometric mean for Model 1a given that samples consisted of 5-6 locations per beach. Each of these variables were natural log-transformed for linearity, mean-centered by unique recruitment-date, then standardized for modelling efficiency and to facilitate setting of priors. The predictive accuracy of the different model specifications was compared using leave-one-out cross-validation ^36^.

The causal effects of the level of water contact and water quality measure variables were evaluated through estimation of marginal effects and contrasts ^37^. Specifically, posterior probability distributions and their summary measures (median and 95% credible intervals [CrI]) were estimated for these effects from a reference grid of model parameters set at standard or default values to calculate adjusted risk differences and risk ratios (RR) (Table S1). A positive association was defined for water contact effects as any risk difference contrast where at least 80% of the posterior distribution indicated an increased incident risk of >1 case of AGI per 1000 beachgoers. Associations for FIB and water quality measure effects on AGI by level of water contact were plotted and visually assessed. The random effects were ignored in the marginal effects and contrasts to produce population-averaged predictions, except when examining differences in water contact effects by site. FIB and water quality measures were back-transformed to the original scale for these predictions. Predictive effects are illustrated through graphical plots, with numerical posterior distribution summaries presented in supplemental file tables. Missing values were possible primarily for the sociodemographic variables, where there was a “prefer not to answer” option on the questionnaire; other questions were mandatory. A complete case analysis was conducted for all models, where observations with missing values for any of the included covariates were removed from the respective model.

All models were conducted using the “brms” package, which uses Stan and Hamiltonian Marko Chain Monte Carlo (MCMC) sampling for model estimation ^34,38^. Models were implemented with 2000 iterations of MCMC sampling, including 1000 warm-up samples, across each of four chains. Marginal effects were calculated using the “marginaleffects” package ^37^. All models were coded in R (version 4.4.0) implemented through the RStudio interface (version 2024.04.02) ^39^. Model code used in this analysis is publicly available from IY’s GitHub account: https://github.com/iany33/Beach_cohort_AGI. An anonymized version of the dataset will be made publicly available on this repository after all planned analyses are completed as per the protocol ^22^.

### 2.5 Model evaluation and sensitivity analyses

All models were assessed for sampling convergence by examining MCMC chains and R-hat values for each parameter ^32,40^. Additionally, posterior probability checks were conducted to determine whether the models were compatible with the observed data ^40^. Specifically, pool adjacent violator (PAV)-adjusted calibration plots were assessed for binary outcomes ^41^. A series of sensitivity analyses were conducted to evaluate the following assumptions and alternate definitions: 1) exposure as a continuous measure of time spent in the water (in min) instead of level of water contact; 2) outcome as diarrhea only instead of AGI; 3) outcome restricted to occurring with three and five days of follow-up compared to seven days; and 4) using weaker (less informative) priors for the water contact and beta parameters. A “negative control analysis” was conducted by running a model containing only the unexposed beachgoers (i.e., those without any water contact) to examine the association with FIB and AGI to identify possibly residual confounding or differential outcome reporting bias ^42,43^. In a deviation from the analysis protocol, a separate random effect was not included in the models to account for some participants being clustered within the same household. Initial models that included this parameter had poor calibration and predictive accuracy, which was likely due to the large number of observations (50.0%) that consisted of a household of only one participant. Instead, a binary covariate was included in each model to indicate if participants were members of a household that had at least one other participant in the study. Additionally, results were compared to an alternative model with one randomly selected participant per household in a sensitivity analysis.

## 3. Results

### 3.1 Descriptive results

A total of 4085 individual beachgoers were recruited from 2730 households. The response proportion for the follow-up survey was 67.6% (n = 2760). Only 66 (1.6%) individuals participated in the study more than once. The sociodemographic characteristics of the recruited beachgoers, tabulations for other confounding variables, and a comparison of characteristics between those who did and did not complete the follow-up survey are shown in Table S2. Approximately one-third of participants (33.6%) reported no water contact, while 22.0% engaged in minimal contact, 28.9% in body immersion, and 15.6% in water contact that resulted in swallowing water (Table S2). Among those who completed the follow-up survey, 73 participants (2.6%) that did not have AGI symptoms at baseline reported AGI illness following their beach visit. Fourteen (19.2%) of these cases were clustered within a household, with two households reporting three cases each and four households reporting two cases each. A summary of the FIB and water quality measures on recruitment days is shown in Table 1.

**Table 1:**
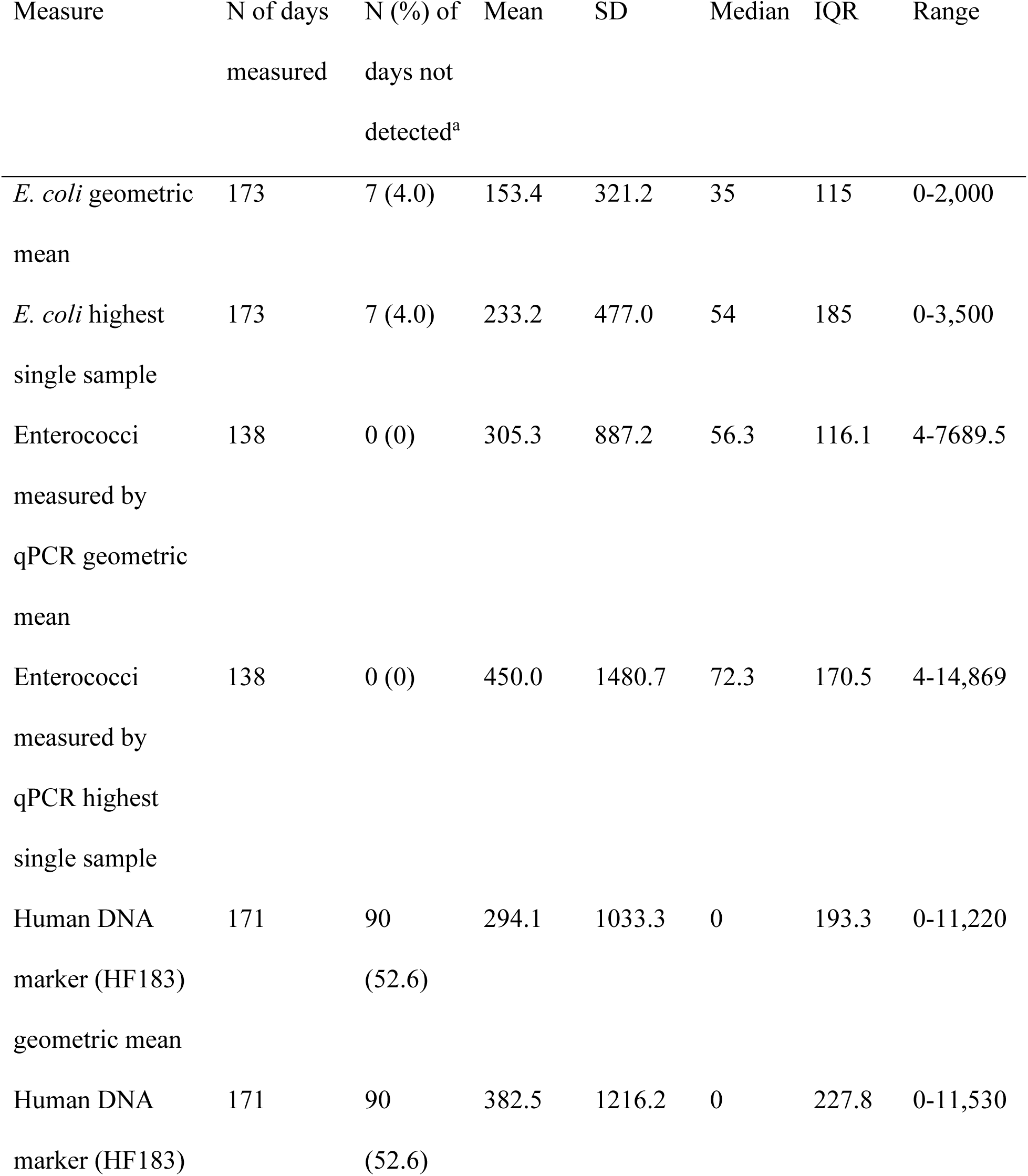
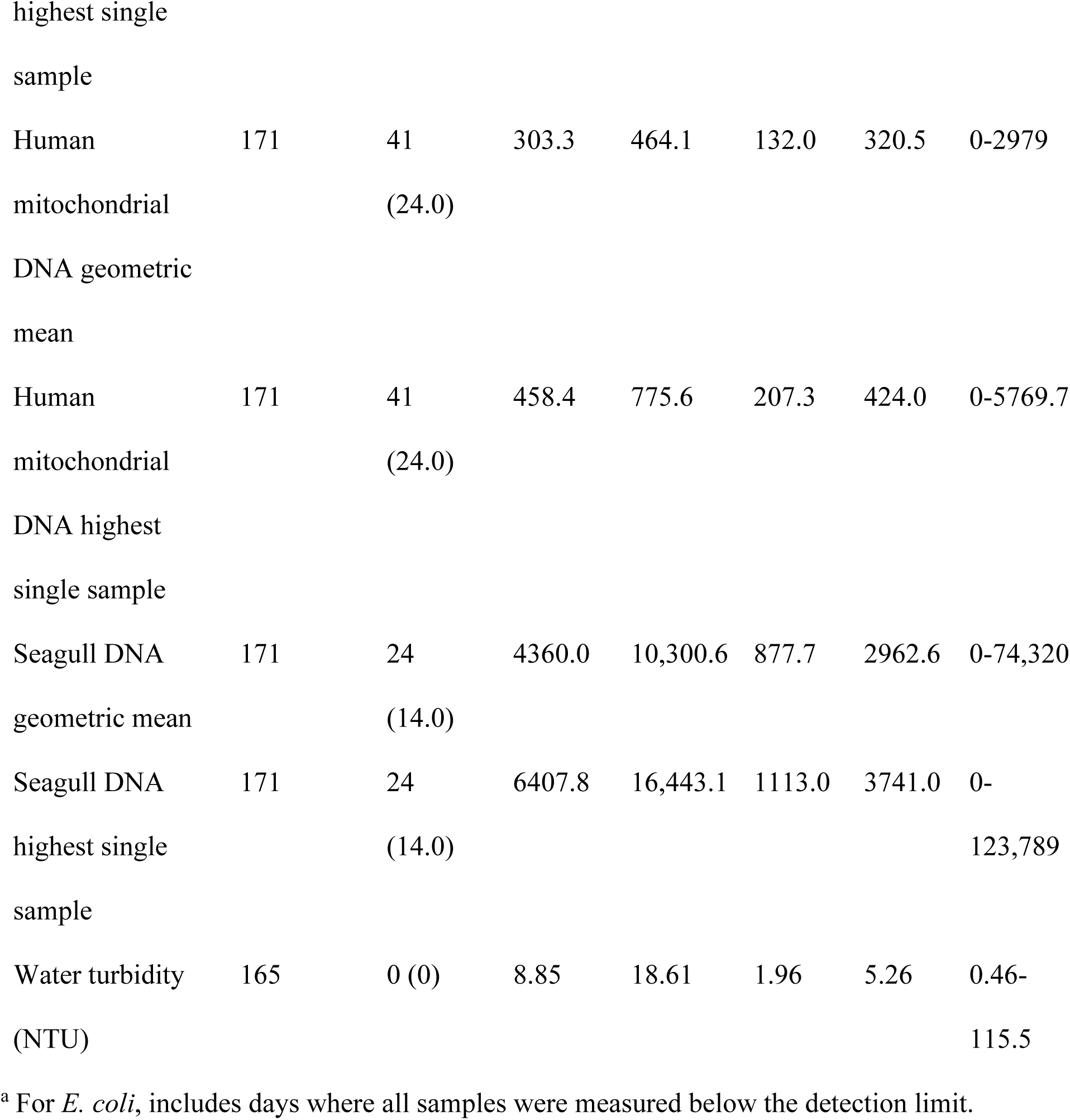
Summary of fecal indicator bacteria (FIB) and water quality measures on recruitment days during the cohort study, 2023-2025.

### 3.2 Relationship between water contact level and AGI

Model 1b with the highest single-sample *E. coli* value as the FIB measure was used to calculate adjusted estimates of the effect of water contact level on AGI incident risk. This model had almost identical predictive accuracy and required fewer assumptions than the *E. coli* geometric mean model (Table S3). A summary of the parameter distributions and MCMC sampling chains from this model are shown in Figure S2, with the PAV-adjusted calibration plot in Figure S3. These diagnostics indicate that the model sampling converged well and had reliable predictive capabilities. The predicted incident risk of AGI per 1000 beachgoers stratified by level of water contact is shown in Figure 1, while the contrast of each level of water contact compared to no contact is shown in Figure 2. These findings indicate that a median of 33 incident cases (95% CrI: 12, 95) of AGI could be expected per 1000 beachgoers that swallow beach water, but with substantial uncertainty in this estimate. There was less uncertainty in the predicted incident risks of the other levels of water contact (Figure 1, Table S4). The risk difference contrasts suggest a dose-response relationship with water contact level (Figure 2). A median estimate of 20 excess cases (95% CrI: 4, 64) of AGI per 1000 beachgoers was predicted for those who swallow water compared to those that visit the beach but have no water contact (Figure 2a), with a RR of 2.62 (95% CrI: 1.31, 5.27) (Figure 2b). There was also substantial uncertainty in these contrasts (Figure 2, Table S4). For those that immerse their body in beach water but do not swallow water, there was a predicted excess of 5 cases (95% CrI: 1, 19) per 1000 beachgoers compared to having no water contact (Figure 2a, Table S4), with a RR of 1.41 (95% CrI: 1.07, 2.26). Minimal water contact was predicted to have very little increase in incident cases compared to no water contact (Figure 2). The probability that these water contact risk differences would exceed specific cut-point values (0, 1, 5, 10, and 20 cases per 1000 beachgoers) is shown in Table S5.

**Figure 1:**
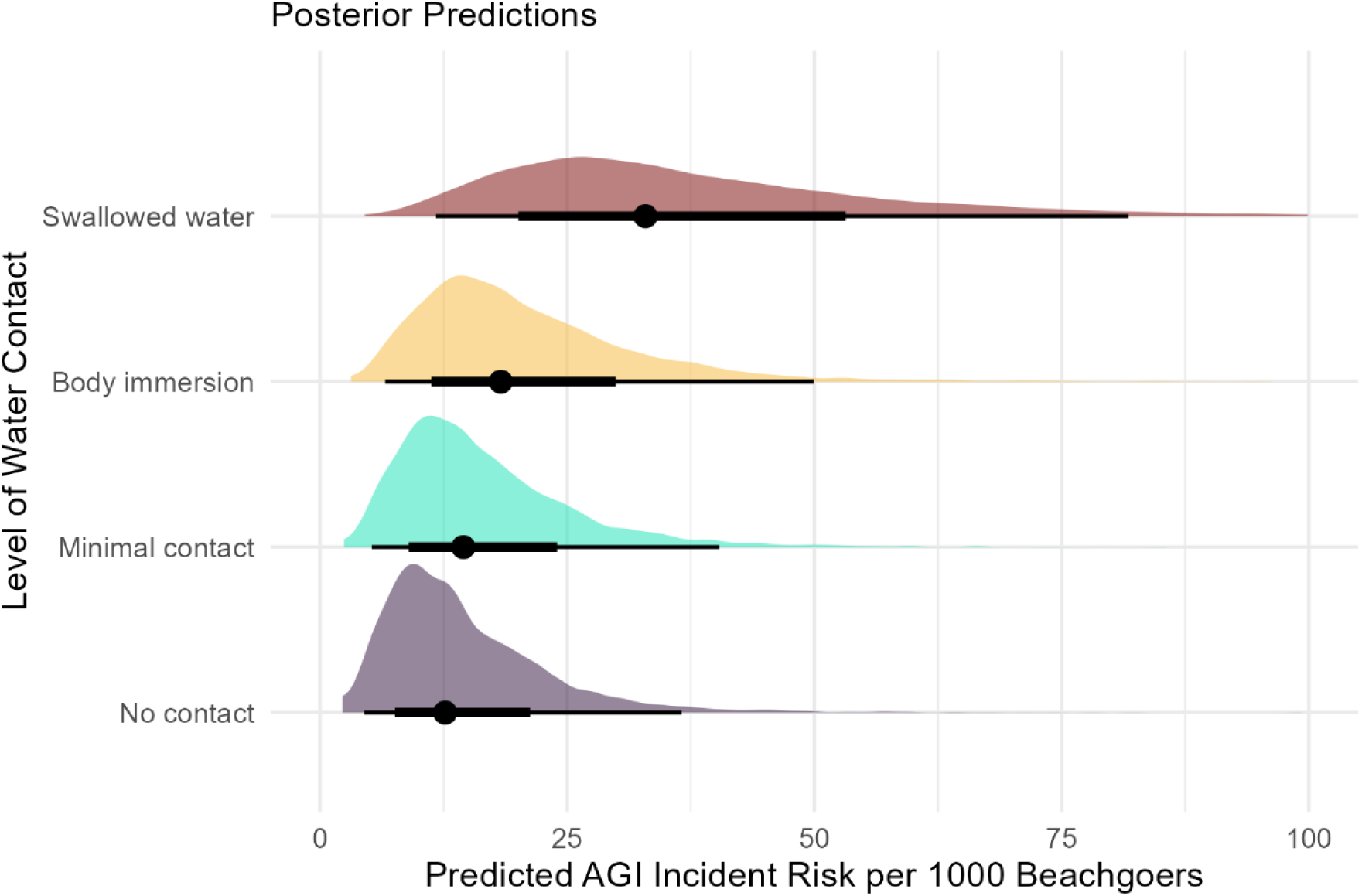
Predicted incident risk of AGI per 1000 beachgoers stratified by level of water contact. Each posterior distribution is summarized at the bottom by its median and 66% and 95% quantile intervals.

**Figure 2:**
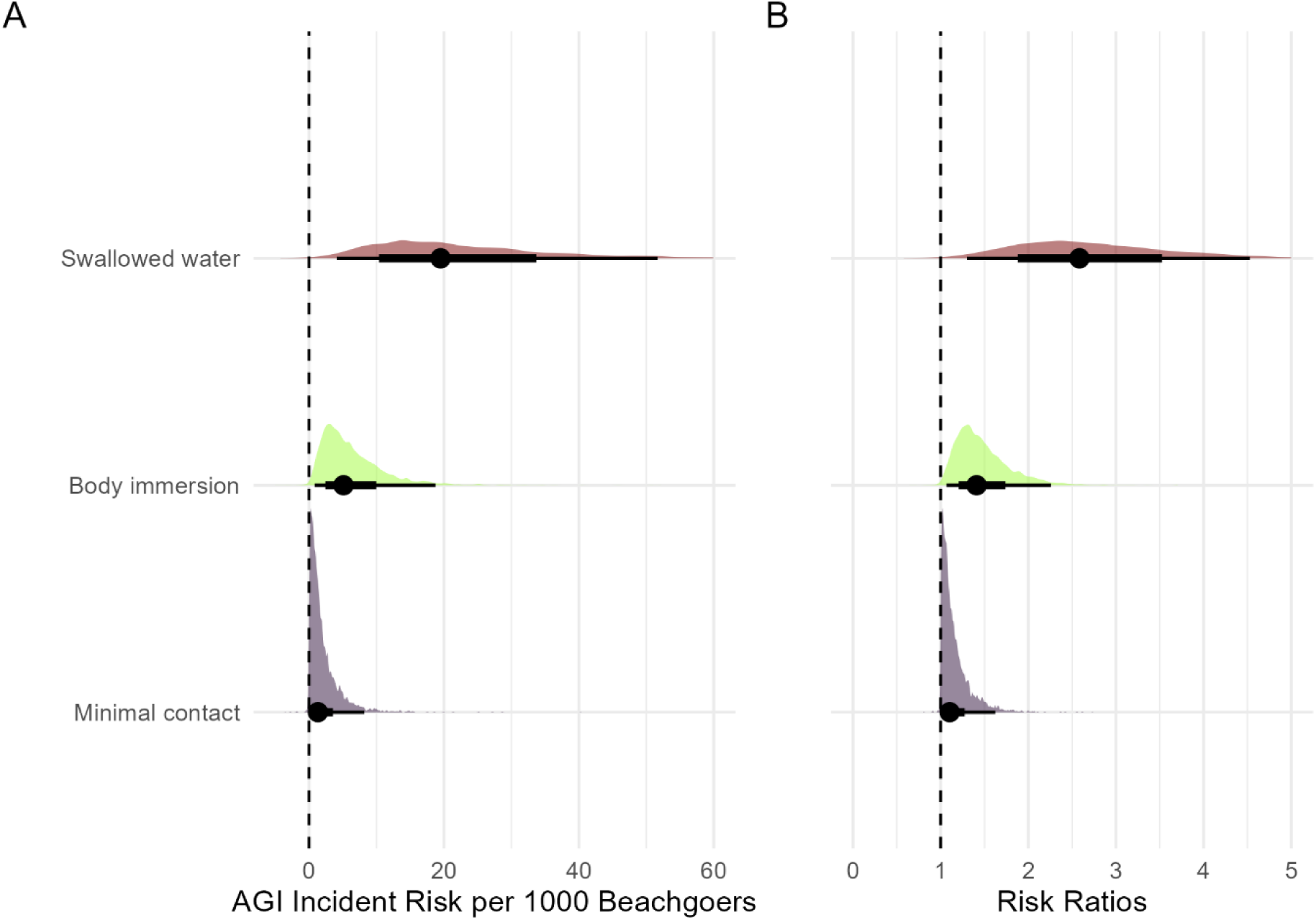
A) Contrasts of the predicted effects of each level of water contact compared to no water contact on the incident risk of AGI per 1000 beachgoers. B) Risk ratios of the predicted effects of each level of water contact compared to no water contact. Each posterior distribution is summarized at the bottom by its median and 66% and 95% quantile intervals.

Stratified contrasts of the effect of water contact level on the incident risk of AGI were calculated for each gender and age category. Gender-specific predictions are shown in Figure 3 and Table S6. The predicted increase in incident AGI risk was slightly lower among those who identified as men or boys compared to women and girls for those that reported swallowing water (18 excess cases [95% CrI: 4, 53] vs. 24 excess cases [95% CrI: 5, 71], respectively). Age-specific contrasts are shown in Figure 4 and Table S6. For the effects of swallowing water and body immersion compared to no water contact, predicted estimates of incident risk of AGI were highest in the 15-19 followed by 0-4 age group, but with substantial uncertainty in these effects across most age groups (Figure 4 and Table S6). Site-specific contrasts are shown in Figure S4 and Table S6 and indicate that predicted effects of water contact were largely consistent across the sites included in this study.

**Figure 3:**
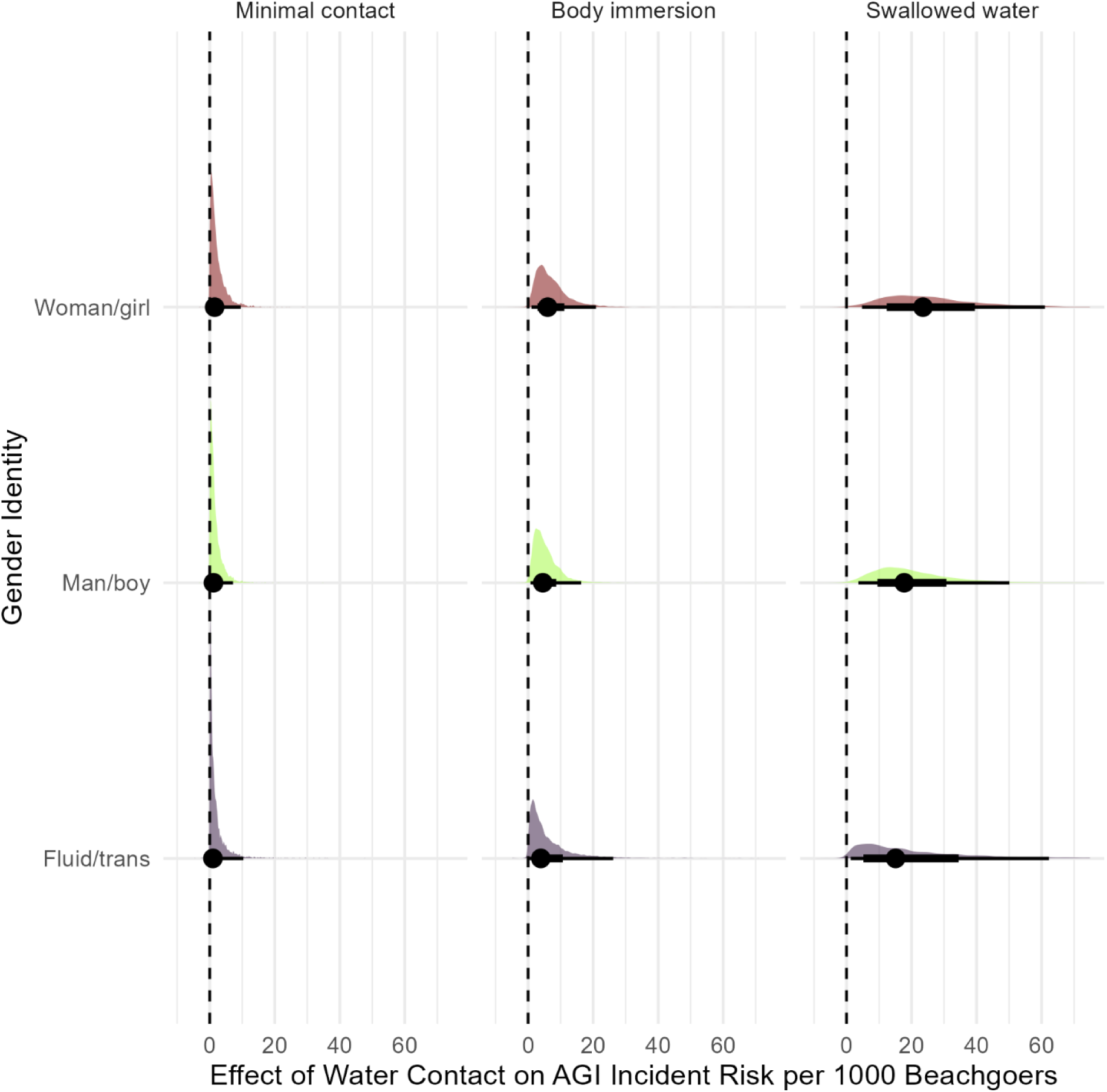
Gender-specific predicted incident risks of AGI per 1000 beachgoers stratified by level of water contact. Each posterior distribution is summarized at the bottom by its median and 66% and 95% quantile intervals.

**Figure 4:**
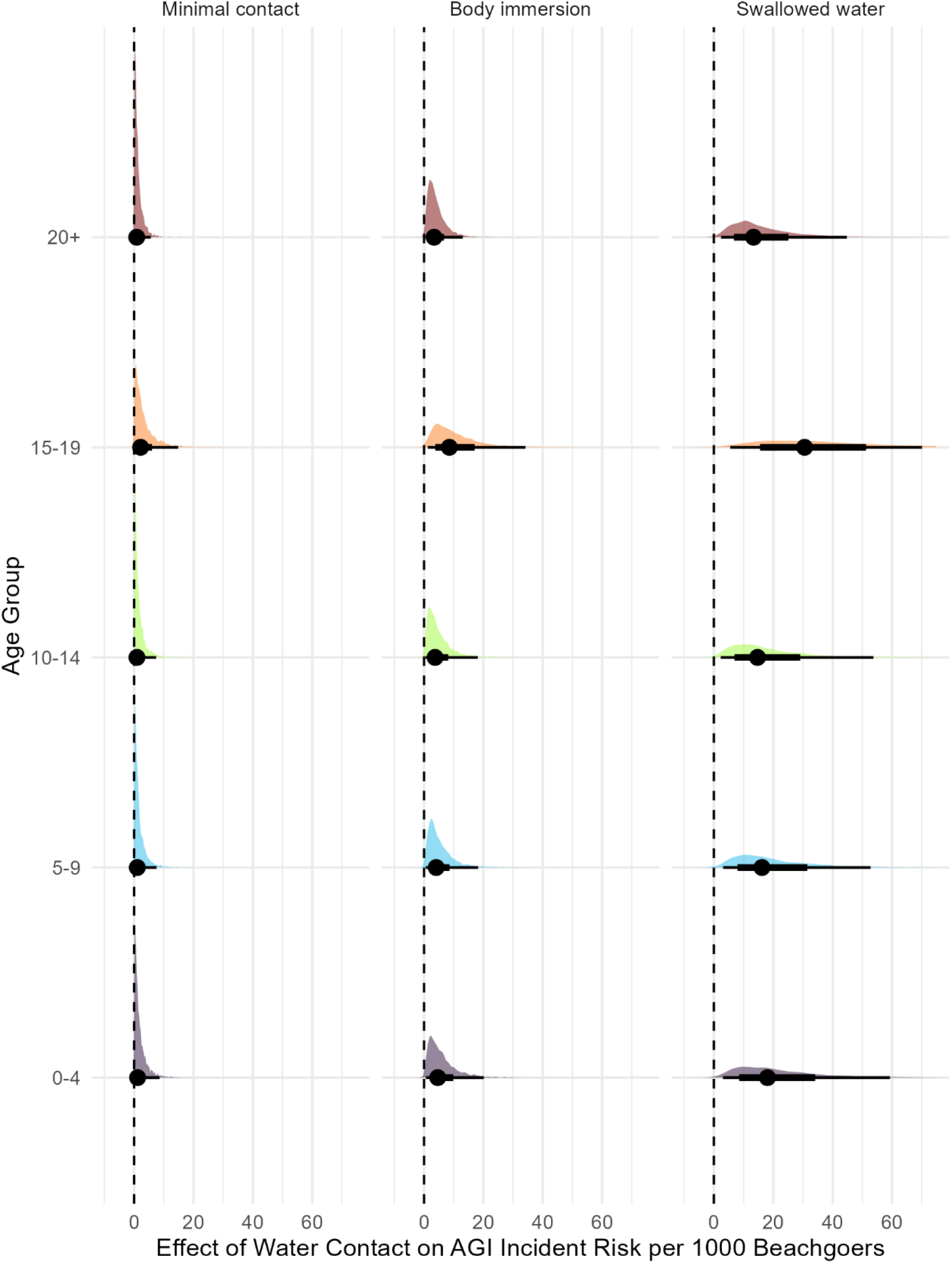
Age-specific predicted incident risks of AGI per 1000 beachgoers stratified by level of water contact. Each posterior distribution is summarized at the bottom by its median and 66% and 95% quantile intervals.

### 3.3 Relationship between different FIB and water quality measures, water contact, and AGI

The predicted effect of each of the different FIB and water quality measures (i.e., Models 1-5) on the incident risk of AGI stratified by level of water contact is shown in Figure 5 and Table S4. In all cases, the highest single sample formulation of the models was used given nearly identical fit to the versions using mean values (Table S3). Higher *E. coli* levels were associated with a slightly increased incident risk of AGI among those who reported body immersion, with a stronger relationship noted among those who swallowed water, though there was considerable uncertainty in these predicted effects (Figure 5). A similar effect was noted for the seagull DNA marker (Figure 5). The human mitochondrial DNA marker showed a weak potential relationship with AGI only among those who swallowed water, while the human HF183/BacR287 marker showed a negative relationship (Figure 5). No association with AGI was noted for the qPCR enterococci and water turbidity measures (Figure 5).

**Figure 5:**
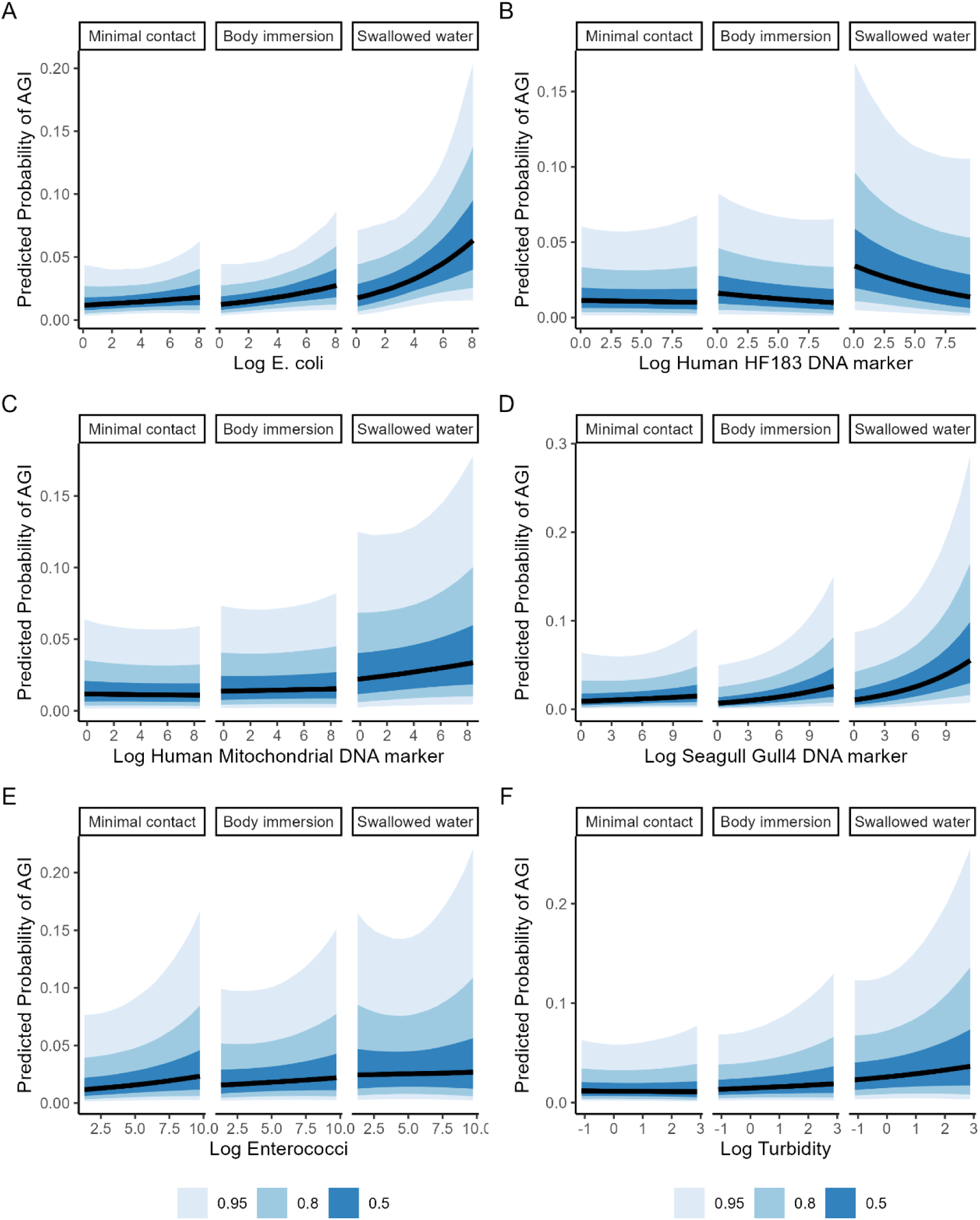
Effects of each of the different FIB and water quality measures on the predicted probability of AGI by level of water contact. A) Culture-based *E. coli*; B) Human HF183 DNA marker; C) Human mitochondrial DNA marker; D) Seagull feces Gull4 DNA marker; E) Enterococci measured through qPCR; F) Water turbidity. Effects are plotted on the log scale.

When examining posterior distributions of effects at specific *E. coli* cut-points (25^th^, 50^th^, 75^th^ and 95^th^ percentiles) stratified by level of water contact, an increased incident risk of AGI was noted at the 50^th^ percentile (∼59 CFU / 100 mL) and above among those who reported body immersion and that reported swallowing water (Figure 6). The magnitude of effect on AGI incident risk and level of uncertainty in estimates was much higher among those who swallowed water (Figure 6). Among those who reported body immersion, incident risks of AGI increased by a median of 7 (95% CrI: 1, 26) and 10 (95% CrI: −2, 41) cases per 1000 beachgoers at 75^th^ and 95^th^ percentile levels, respectively. Among those who swallowed water compared to those who had no water contact, median increases in incident risks were 27 (95% CrI: 6, 83) and 38 (95% CrI: 3, 126) cases per 1000 beachgoers at 75^th^ and 95^th^ percentile levels, respectively.

**Figure 6:**
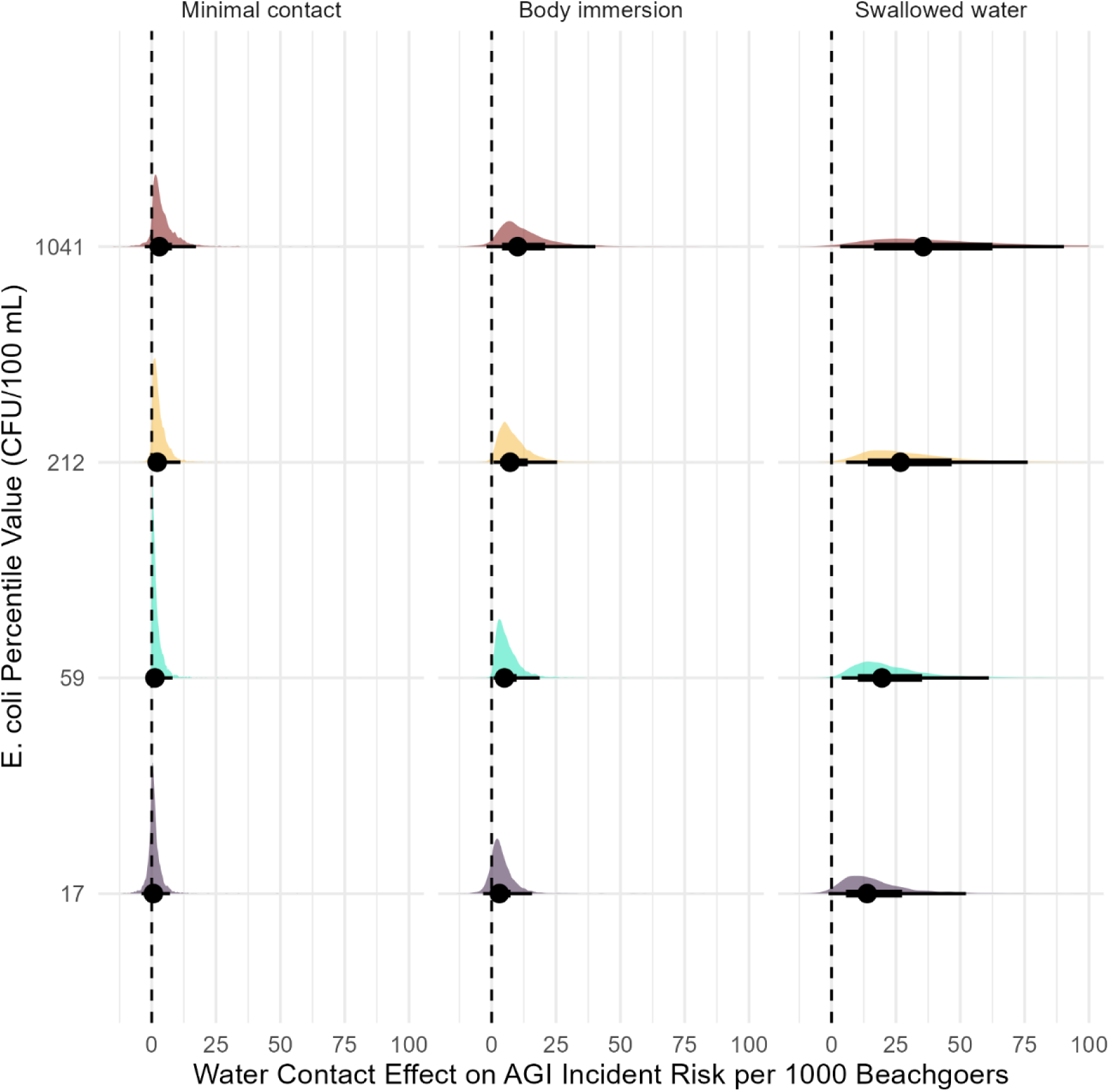
Predicted effects of *E. coli* on AGI incident risk per 1000 beachgoers by level of water contact (compared to no water contact) at four specific percentile values (25^th^, 50^th^, 75^th^, and 95^th^). Each posterior distribution is summarized at the bottom by its median and 66% and 95% quantile intervals.

### 3.4 Sensitivity analysis results

Results of each of the sensitivity analyses are shown in Table S7 and Figures S5-6. All are compared to Model 1b (*E. coli* as FIB with highest single sample value). In the model examining time in the water in minutes as a continuous exposure variable, a slight positive relationship was noted with AGI as time in the water increased (Table S7 and Figure S5). Only 57 (2.1%) follow-up participants specifically reported experiencing diarrhea after their beach visit. In the model examining this outcome instead of AGI, a stronger relationship was noted for the effects of swallowing water and body immersion compared to no contact on the RR scale, with an attenuated relationship on the risk difference scale and for the effect of *E. coli* (Table S7 and Figure S5). Reducing the follow-up period to 3- and 5-days reduced the water contact effects, particularly for the 3-day follow-up, with similar effects on the relationship between *E. coli* levels and AGI (Table S7 and Figure S5). The model with weaker priors on parameters slightly increased the RR effects for the different levels of water contact, while the *E. coli* effect was attenuated with increased uncertainty (Table S7 and Figure S5). The model with one randomly selected participant per household had similar results to Model 1b, with slightly increased effects on the risk difference scale and attenuated effects on the risk ratio scale for water contact and increased uncertainty for the *E. coli* effect (Table S7 and Figure S5). The negative control analysis model indicated no relationship between *E. coli* levels and AGI among respondents that had no water contact (Figure S6).

## 4. Discussion

This study reports on the first large, Canadian cohort study of recreational water illness due to beach water contact. The FIB levels during the recruitment days were largely well below current BAV levels, which is consistent with no obvious point sources of fecal pollution at the included beaches. Therefore, the results of this study are most comparable to other epidemiological studies at non-point source affected beaches ^44^. The low incident risks of AGI in this study, compared to prior U.S. and European studies ^19,44,45^, is likely due to the low levels of fecal pollution identified. This occurred despite selection of beaches that were anticipated to have fluctuations in FIB levels while also being popular enough to support recruitment efforts. These findings may be indicative of strong beach management programs across the sites for public health and tourism-related reasons. For example, beaches that are known to public health authorities and operators as being high-risk for fecal contamination are likely to receive more attention for remediation efforts, or to not be recommended as public bathing sites ^46,47^.

This study used an ordinal dose-response exposure variable to assess risk of AGI from increasing levels of water contact. Swallowing water, in particular, was associated with an increased AGI incident risk compared to those who had no water contact. The incident risk estimates are comparable to those found in a recent quantitative microbial risk assessment (QMRA) for recreation in Ontario freshwater sources (∼1-37 cases per 1000 beachgoers) ^48^. A slight positive relationship with AGI incident risk was also noted in the sensitivity analysis that considered the total amount of time in the water (in min) as an alternative exposure definition, suggesting that this effect is robust to different ways of measuring water exposure. Given the large difference in risk when swallowing water compared to no water contact, future public health risk communication might focus specifically on informing beachgoers of risks of this behaviour in addition to swimming or body immersion. However, it can be difficult for beachgoers to prevent the possibility of swallowing water when engaging in water activities, especially for children ^49^.

This is one of the first beach cohort studies to report both gender- and age-stratified AGI risk estimates. A slightly increased risk of AGI was found among women compared to men, which corresponds with previous research examining AGI in Canada ^50,51^. This gender difference could be related to behavioural factors such as higher levels of interaction among women with young children, potential recall or reporting biases (e.g., women could be more likely to report AGI than men), or sex-based biological differences in gastrointestinal pathophysiology or susceptibility ^50,52^. Previous studies have found that AGI risks due to water contact are highest among children aged 10 or younger ^3,4^. However, results of this study suggested that teenagers aged 15-19 years old were at greatest risk. The reasons for this increased risk are not clear but could be due to reporting or recall biases or potentially more intense water behaviours in this age group among participants in this study. Age categories were predetermined in the study protocol, but the risk estimates are very uncertain given the small sample sizes with these groups across the levels of water contact.

Culture-based *E. coli* had the strongest positive association with AGI among the FIB and water quality measures evaluated, a relationship that was much stronger among participants that reported swallowing water compared to no water contact. Globally, researchers studying beaches affected by non-point sources of fecal pollution have not identified a single FIB or water quality measure as a consistent predictor of AGI ^44^. FIB measures are also not consistently predictive of waterborne pathogens that can cause AGI ^53^. However, current BAV levels in Canada and the US are based on both *E. coli* and qPCR enterococci measures, with the former being the most commonly used by public health authorities for routine beach water monitoring across Canada ^16^.

There is increasing momentum for public health laboratories to adopt rapid-measured FIB measures for beach water management rather than relying on traditional culture-based methods to facilitate more timely and responsive public health notifications and warnings ^54,55^. It is possible that the lack of relationship noted for enterococci in this study is at least partially explained by lack of power given that water samples were not tested for this measure in the 2023 recruitment season in Toronto. Rapid qPCR methods have also been developed for *E. coli* in beach water, which should also be explored in future studies within a Canadian context ^56^. Water turbidity was also evaluated as another potential rapid indicator of AGI risks given its relationship to FIB in beach water and potential association with AGI in drinking water in epidemiological studies ^57,58^. However, no evidence of a relationship was found, suggesting that water turbidity is not a reliable indicator of AGI risks from beach water exposures in Canada. Overall, the results indicate that despite limitations in turnaround time of results, culture-based *E. coli* are still a good indicator of AGI risks at the beaches in this study.

Three MST DNA markers were evaluated in this study, two different human indicators and a seagull feces marker. Previous research of human-associated *Bacteroides* markers, including HF183 as well as other markers, have also shown inconsistent or no relationship with AGI among beachgoers ^59,60^. The human mitochondrial marker reflects a broader impact of human activity than HF183 because it could be present due to shedding of human skin cells, hair, and saliva, as well as fecal matter, while HF183 is a specific marker of human fecal contamination ^29,61^. Human-associated DNA markers may be less persistent in beach water than FIB, and thus their presence may be more useful as an indicator of recent contamination events rather than human illness risks ^59^. Seagull feces contamination was evident across most beaches and was likely a main contributor to overall *E. coli* levels. Seagull feces can harbour bacterial pathogens such as *Campylobacter* and *Salmonella*, which could explain the positive association with AGI identified in this study ^62,63^. These results are also consistent with a previous case-control study of sporadic campylobacteriosis cases in Canada, which identified swimming in natural water bodies as a risk factor for illness ^64^. Results suggest that human MST markers were not useful as FIB measures for human health risk at the beaches in this study, and they likely have more utility as part of beach sanitary surveys and targeted monitoring to identify sources of recent contamination and contributors to sewage pollution at beaches, which can guide pollution prevention efforts.

The models predicted that incident risks of AGI are expected to increase on average among those who report body immersion and swallowing water compared to those who had no water contact, even at *E. coli* contamination levels at the 50^th^ percentile and above as observed in this study (∼59 CFU/100mL and higher). The average increase in risk was marginal for the body immersion group. Among those who reported swallowing water, median incident risks were higher, but with substantial uncertainty. The current Health Canada BAV for *E. coli* is 235 CFU/100mL, which corresponds to an increase in risk of 36 illnesses per 1000 exposed beachgoers and is based on the 75^th^ percentile of FIB levels from previous US EPA beach cohort studies ^16^. Results suggest that, on average, a similar increase in risk of AGI can be expected to occur at higher levels of *E. coli* pollution when individuals swallow water (see Figure 6). Given the considerable uncertainty in predicted risk estimates, beach managers should aim to keep FIB levels low and ideally less than the 75^th^ percentile level in this study to lower population illness risks, particularly among vulnerable groups such as young children who have limited ability to control their high-risk behaviours (e.g., swallowing water).

The sensitivity analysis using diarrhea as the outcome resulted in stronger relative associations for swallowing water and body immersion vs. no water contact when compared to the broader definition of AGI, which is similar to prior cohort studies ^3,24^. Fewer participants reported diarrhea specifically in this study, likely contributing to the lower risk differences observed for the effect of water contact on this outcome when compared to AGI. However, in contrast with previous studies, shorter follow-up times attenuated the relationships with AGI ^23,24^. This finding could be due to differences in etiology of AGI in this study, which might have been caused by bacterial or protozoan pathogens compared to enteric viruses, particularly norovirus, which have a shorter incubation period ^65^. For example, previous research in Lake Ontario, near beaches at the Toronto sites in this study, did not find culturable enteric viruses during the swimming season, while *Campylobacter*, *Cryptosporidium*, and *Giardia* were sporadically found in the summer ^66^. Future research to evaluate possible pathogens causing AGI among beachgoers in Canada is needed.

There are some limitations to this study. Over three years of data collection at five different sites across Canada, a similar number of participants were recruited as previous prospective cohort studies, but fewer than the cohort studies conducted in the U.S. in the early 2000s ^3,4,44,67^. While a gift card incentive was offered along with other approaches (e.g., multiple reminders and ways to respond) to encourage completion of the follow-up survey, approximately one-third of participants did not complete the follow-up. Characteristics of those who did not complete the follow-up survey were largely similar to those who did, but the possibility of differences in AGI risks between these two groups cannot be ruled out.

Given that AGI was self-reported, it is possible that some participants selectively reported illness following their beach visit based on their level of water contact. However, the negative control analysis found that there was no association between FIB levels and AGI among those who visited the beaches in this study and did not have any water contact, suggesting that the water contact-FIB relationship with AGI may have been robust to possible differential misclassification bias. Including a more objective measure of illness would be ideal, such as salivary immunoassays that can detect multiple potential enteric pathogens, which could complement the non-specific symptom based case definitions used in this and previous studies^68,69^. However, such an approach would likely make participant recruitment even more challenging; beachgoers approached during recruitment in this study were already reluctant to participate and be disturbed during their beach visit to complete questionnaires, especially those with young children, even with a gift card incentive provided ^22^. Another potential limitation is that different laboratories were used in each province to test for *E. coli*, with membrane filtration used in Ontario and Vancouver, and Colilert used in the other two provinces. However, previous research has found that these methods are comparable ^70^, and site variability was accounted for in the models.

## 5. Conclusion

In a large, prospective cohort study at non-point source affected beaches in Canada, a low overall incidence of AGI was identified. However, swallowing water, and to a lesser extent body immersion, were associated with increased risks of illness compared to no water contact, though with substantial uncertainty in the predicted magnitude of effects. Gender- and age-specific differences in effects were identified and should be further investigated to inform public health preventative outreach and messaging strategies. *Escherichia coli* and seagull feces DNA marker levels were associated with AGI. AGI risks were predicted to increase when *E. coli* was at the median or higher levels of contamination among those who immersed their body in the water or swallowed water. Since this study only examined AGI symptoms among participants, future research is needed to investigate the enteric pathogens associated with AGI at beaches with non-point sources of fecal pollution. Public health agencies responsible for beach water quality management should consider integrating level of water contact into beach advisory messaging and advisories.

## Supporting Information

Supporting Information: Additional study analytical details, numerical results, and sensitivity analysis results as Tables S1-S7 and Figures S1-S6 (DOCX).

## Data Availability

All data produced in the present study are available upon reasonable request to the authors

https://github.com/iany33/Beach_cohort_AGI

## Acknowledgements

This work was supported by the Canadian Institutes of Health Research (CIHR), grant numbers PJT 185894 and PJT 192023 (P.I. Young). We thank all the beachgoers that agreed to participate in this study. We would also like to thank all of the student data collectors at each of our sites across Canada for their efforts in participant recruitment and enthusiasm towards this project.

## Author contributions: CRediT

**Ian Young:** Conceptualization, Methodology, Investigation, Formal analysis, Visualization, Supervision, Project administration, Funding acquisition, Writing - Original Draft. **Rachel Jardine, Binyam N. Desta, J. Johanna Sanchez, Jordan Tustin**: Conceptualization, Methodology, Investigation, Supervision, Project administration, Funding acquisition, Writing - Review & Editing. **Thomas A. Edge, Herb E. Schellhorn:** Conceptualization, Methodology, Investigation, Resources, Writing - Review & Editing. **Shannon E. Majowicz, David L. Pearl, Sarah Elton, Andrea Nesbitt, Teresa Brooks**: Conceptualization, Methodology, Writing - Review & Editing. **Michael Schwandt, Dylan Lyng, Brandon Krupa, Elizabeth Montgomery, Mahesh Patel**: Conceptualization, Project administration, Writing - Review & Editing.

