## Supplemental File for "The associations between recreational water contact, water quality measures, and acute gastrointestinal illness among Canadian beachgoers: a prospective cohort study"

**Supporting Information:**

Table S1: Reference grid used for marginal effects calculations for regression models reported in this article

| Variable | Referent category(s) for predictions |
|---|---|
| FIB or water quality measure (continuous, mean-centered and standardized) | Mean value for water contact predictions (grouped by unique location-recruitment date); sequence across range of values across the location-recruitment dates for FIB/water quality predictions |
| Age | Overall predictions averaged across all categories; age-specific predictions for each category |
| Gender | Overall predictions averaged across all categories; gender-specific predictions for each category |
| Education | Bachelor's degree* |
| Presence of GI condition | No* |
| Immune-compromised | No* |
| Presence of allergies | No* |
| Engaged in other recreational water activities in prior 2 weeks | Yes* |
| Consumed food at the beach | Yes* |
| Played with sand at the beach | No* |

| | |
|---|---|
| Was member of a household where at least one other person also participated in the study | Yes* |

* Selected for predictions as these were the most commonly reported values for each variable.

Table S2: Description tabulation of sociodemographic and confounding variables for participants in the study (with variable categories as used in analytical models)

| Variable | N (%) total participants | N (%) that completed follow-up | N (%) that did not complete follow-up |
|---|---|---|---|
| Age group | | | |
| 0-4 | 160 (3.9) | 108 (3.9) | 52 (4.0) |
| 5-9 | 379 (9.3) | 249 (9.0) | 130 (9.9) |
| 10-14 | 321 (7.9) | 189 (6.9) | 132 (10.1) |
| 15-19 | 659 (16.2) | 412 (15.0) | 247 (18.9) |
| 20+ | 2,540 (62.6) | 1,794 (65.2) | 746 (57.1) |
| Prefer not to answer | 26 | 8 | 18 |
| Gender | | | |
| Woman/girl | 2,392 (59.3) | 1,677 (61.4) | 715 (54.9) |
| Man/boy | 1,582 (39.2) | 1,014 (37.1) | 568 (43.6) |
| Fluid/trans | 60 (1.5) | 41 (1.5) | 19 (1.5) |
| Prefer not to answer | 51 | 28 | 23 |
| Education (highest in household) | | | |
| High school or less | 655 (16.9) | 397 (15.0) | 258 (21.1) |
| College/trades | 760 (19.6) | 499 (18.8) | 261 (21.4) |
| Bachelors | 1,120 (28.9) | 812 (30.6) | 308 (25.2) |
| Post-graduate | 1,339 (34.6) | 944 (35.6) | 395 (32.3) |
| Prefer not to answer | 211 | 108 | 103 |
| Water contact level | | | |
| No contact | 1359 (33.2) | 926 (33.6) | 433 (32.7) |
| Minimal contact | 858 (21.0) | 606 (22.0) | 252 (19.0) |

| | | | |
|---|---|---|---|
| Body immersion | 1235 (30.2) | 798 (28.9) | 437 (33.0) |
| Swallowed water | 633 (15.5) | 430 (15.6) | 203 (15.3) |
| Presence of GI condition | | | |
| No | 3,949 (96.7) | 2,660 (96.4) | 1,289 (97.3) |
| Yes | 136 (3.3) | 100 (3.6) | 36 (2.7) |
| Immune-compromised | | | |
| No | 4,000 (97.9) | 2,699 (97.8) | 1,301 (98.2) |
| Yes | 85 (2.1) | 61 (2.2) | 24 (1.8) |
| Presence of allergies | | | |
| No | 3,680 (90.1) | 2,463 (89.2) | 1,217 (91.8) |
| Yes | 405 (9.9) | 297 (10.8) | 108 (8.2) |
| Engaged in other recreational water activities in prior 2 weeks | | | |
| No | 1,449 (35.5) | 893 (32.4) | 556 (42.0) |
| Yes | 2,636 (64.5) | 1,867 (67.6) | 769 (58.0) |
| Consumed food at the beach | | | |
| No | 1,725 (42.2) | 1,136 (41.2) | 589 (44.5) |
| Yes | 2,360 (57.8) | 1,624 (58.8) | 736 (55.5) |
| Played with sand at the beach | | | |
| No | 2,108 (51.6) | 1,435 (52.0) | 673 (50.8) |
| Yes | 1,977 (48.4) | 1,325 (48.0) | 652 (49.2) |
| Was member of a household where at least one other person also participated in the study | | | |
| No | 1,860 (45.5) | 1,262 (45.7) | 598 (45.1) |
| Yes | 2,225 (54.5) | 1,498 (54.3) | 727 (54.9) |

| Number of participants per household | | | |
|---|---|---|---|
| 1 | 1,860 (45.5) | 1,262 (45.7) | 598 (45.1) |
| 2 | 1,092 (26.7) | 723 (26.2) | 368 (27.8) |
| 3 | 597 (14.6) | 406 (14.7) | 191 (14.4) |
| 4 | 376 (9.2) | 262 (9.5) | 114 (8.6) |
| 5 | 130 (3.2) | 87 (3.2) | 43 (3.2) |
| 6 | 30 (0.7) | 20 (0.7) | 10 (0.8) |

Table S3: Comparison of predictive accuracy of alternative models

| Model | N observations | Difference in expected log predictive density[a] | SE in difference |
|---|---|---|---|
| *E. coli* model comparison | 2638 | | |
| Model 1a (mean) | | -0.1 | 0.4 |
| Model 1b (highest single sample) | | 0 | 0 |
| Human DNA marker (HF183) model comparison | 2612 | | |
| Model 2a (mean) | | 0 | 0 |
| Model 2b (highest single sample) | | -0.2 | 0.3 |
| Human mitochondrial DNA marker model comparison | 2612 | | |
| Model 3a (mean) | | 0 | 0 |
| Model 3b (highest single sample) | | -0.4 | 0.3 |
| Seagull feces DNA marker model comparison | 2612 | | |
| Model 4a (mean) | | 0 | 0 |
| Model 4b (highest single sample) | | -0.2 | 0.3 |
| Enterococci model comparison | 2163 | | |
| Model 5a (mean) | | 0 | 0 |
| Model 5b (highest single sample) | | 0 | 0.4 |

[a] Best-fitting model for each comparison is noted with a 0 expected log predictive density. Note that comparisons were only possible and conducted for models with the same number of observations.

Table S4: Summary of posterior distributions of predicted effects and contrasts of models reported in this article (predictions and contrasts are per 1000 beachgoers)

| **Parameter / contrast** | **Median posterior estimate (95% credible interval)** | | | | | |
|---|---|---|---|---|---|---|
| | **Model 1b: *E. coli* highest single sample** | **Model 2b: Human DNA marker (HF183) highest single sample** | **Model 3b: Human mitochondrial DNA marker highest single sample** | **Model 4b: Seagull feces DNA marker highest single sample** | **Model 5b: Enterococci highest single sample** | **Model 6: Turbidity** |
| No. observations | 2638 | 2612 | 2612 | 2612 | 2163 | 2496 |
| Swallowed water | 33 (12, 95) | 31 (11, 93) | 32 (11, 92) | 32 (11, 102) | 30 (9, 107) | 30 (10, 96) |
| Body immersion | 18 (7, 52) | 17 (6, 51) | 17 (6, 51) | 17 (6, 55) | 21 (75, 74) | 17 (6, 53) |
| Minimal contact | 15 (5, 41) | 13 (4, 41) | 13 (5, 41) | 14 (5, 46) | 18 (6, 67) | 14 (5, 43) |
| No water contact | 13 (4, 37) | 11 (4, 37) | 12 (4, 36) | 13 (4, 42) | 17 (5, 64) | 12 (4, 39) |
| Contrast: swallowed water-no contact | 20 (4, 64) | 19 (4, 63) | 20 (4, 66) | 18 (2, 66) | 12 (-4, 60) | 17 (3, 63) |

| | | | | | | |
|---|---|---|---|---|---|---|
| Contrast: body immersion-no contact | 5 (1, 19) | 5 (1, 20) | 5 (1, 20) | 4 (-1, 19) | 4 (-2, 20) | 5 (1, 19) |
| Contrast: minimal contact-no contact | 1 (0, 8) | 1 (0, 9) | 1 (0, 9) | 1 (-1, 9) | 1 (-1, 9) | 1 (0, 8) |
| Risk ratio: swallowed water-no contact | 2.62<br>(1.31, 5.27) | 2.73<br>(1.37, 5.74) | 2.75<br>(1.35, 5.79) | 2.48<br>(1.14, 5.18) | 1.79<br>(0.82, 3.71) | 2.52<br>(1.22, 5.09) |
| Risk ratio: body immersion-no contact | 1.41<br>(1.07, 2.26) | 1.43<br>(1.07, 2.45) | 1.44<br>(1.08, 2.43) | 1.31<br>(0.9, 2.24) | 1.24<br>(0.91, 1.95) | 1.39<br>(1.04, 2.30) |
| Risk ratio: minimal contact-no contact | 1.10<br>(1.00, 1.62) | 1.11<br>(1.00, 1.72) | 1.12<br>(1.00, 1.68) | 1.08<br>(0.92, 1.59) | 1.06<br>(0.98, 1.44) | 1.09<br>(1.00, 1.62) |
| FIB overall effect* | 0.007<br>(-0.003, 0.025) | -0.007<br>(-0.034, 0.008) | 0.003<br>(-0.020, 0.026) | 0.007<br>(-0.002, 0.030) | 0.001<br>(-0.010, 0.015) | 0.002<br>(-0.010, 0.016) |
| FIB:swallowed water interaction* | 0.013<br>(-0.006, 0.051) | -0.015<br>(-0.072, 0.014) | 0.008<br>(-0.033, 0.056) | 0.012<br>(-0.004, 0.055) | 0.001<br>(-0.022, 0.022) | 0.004<br>(-0.019, 0.031) |

| | | | | | | |
|---|---|---|---|---|---|---|
| FIB:body immersion interaction* | 0.005 (-0.004, 0.020) | -0.004 (-0.026, 0.009) | 0.001 (-0.018, 0.020) | 0.005 (-0.002, 0.027) | 0.001 (-0.010, 0.014) | 0.002 (-0.008, 0.015) |
| FIB:minimal contact interaction* | 0.002 (-0.007, 0.013) | -0.001 (-0.015, 0.013) | -0.001 (-0.018, 0.012) | 0.002 (-0.005, 0.015) | 0.002 (-0.006, 0.017) | 0.000 (-0.010, 0.008) |

[*] Effects are on the log transformed, mean-centered and standardized scale and contrast across the interquartile range (IQR) of FIB values

Table S5: Probability that water contact risk difference contrasts would exceed cut-point values of 0, 1, 5, 10, and 20 cases per 1000 beachgoers

| | **Probability of exceeding threshold** | | |
|---|---|---|---|
| **Cut-point values** | **Swallowed water vs. no contact** | **Body immersion vs. no contact** | **Minimal contact vs. no contact** |
| 0 cases per 1000 beachgoers | 0.996 | 0.996 | 0.991 |
| 1 case per 1000 beachgoers | 0.993 | 0.968 | 0.593 |
| 5 cases per 1000 beachgoers | 0.963 | 0.510 | 0.089 |
| 10 cases per 1000 beachgoers | 0.844 | 0.168 | 0.014 |
| 20 cases per 1000 beachgoers | 0.499 | 0.021 | 0.002 |

Table S6: Summary of gender-, age-, and site-stratified posterior distributions of predicted risk difference contrasts (per 1000 beachgoers) for water contact exposure

| | **Median posterior estimate (95% credible interval)** | | |
|---|---|---|---|
| **Variable / category** | **Swallowed water vs. no contact** | **Body immersion vs. no contact** | **Minimal contact vs. no contact** |
| **Gender** | | | |
| Boys/men | 18 (4, 53) | 5 (1, 16) | 1 (0, 7) |
| Girls/women | 24 (5, 71) | 6 (1, 21) | 1 (0, 10) |
| Gender fluid/trans | 16 (1, 91) | 4 (0, 26) | 1 (0, 10) |
| **Age group** | | | |
| 0-4 years | 18 (3, 67) | 5 (1, 20) | 1 (0, 9) |
| 5-9 years | 16 (3, 58) | 4 (1, 18) | 1 (0, 8) |
| 10-14 years | 15 (1, 61) | 4 (1, 18) | 1 (0, 7) |
| 15-19 years | 33 (6, 111) | 9 (1, 34) | 2 (0, 15) |
| 20+ years | 13 (2, 47) | 3 (1, 13) | 1 (0, 6) |
| **Site** | | | |
| Toronto | 19 (4, 60) | 5 (1, 18) | 1 (0, 8) |
| Vancouver | 18 (4, 57) | 5 (1, 18) | 1 (0, 8) |
| Manitoba | 20 (4, 65) | 5 (1, 19) | 1 (0, 8) |
| Halifax | 19 (4, 62) | 5 (1, 19) | 1 (0, 8) |
| Niagara Region | 20 (4, 61) | 5 (1, 19) | 1 (0, 8) |

Table S7: Summary and comparison of predicted posteriors of parameters of interest for sensitivity analysis models (contrasts are per 1000 beachgoers)

| **Parameter / contrast** | **Median posterior estimate (95% credible interval)** | | | | | | |
|---|---|---|---|---|---|---|---|
| | **Model 1b** | **Time in water exposure** | **Diarrhea outcome** | **3-day follow-up** | **5-day follow-up** | **Weaker priors[a]** | **One participant per household** |
| No. observations | 2638 | 2638 | 2638 | 2638 | 2638 | 2638 | 1833 |
| Contrast: swallowed water-no contact | 20 (4, 64) | N/a | 14 (4, 50) | 7 (-3, 40) | 14 (0, 61) | 19 (4, 64) | 31 (3, 100) |
| Contrast: body immersion-no contact | 5 (1, 19) | N/a | 3 (1, 13) | 2 (-1, 13) | 4 (0, 18) | 5 (1, 21) | 8 (1, 30) |
| Contrast: minimal | 1 (0, 8) | N/a | 0 (0, 4) | 1 (0, 5) | 1 (0, 8) | 2 (0, 13) | 2 (0, 13) |

| | | | | | | | |
|---|---|---|---|---|---|---|---|
| contact-no contact | | | | | | | |
| Risk ratio: swallowed water-no contact | 2.62 (1.31, 5.27) | N/a | 3.68 (1.73, 7.76) | 2.11 (0.76, 5.46) | 2.27 (1.00, 4.98) | 2.93 (1.42, 6.36) | 2.30 (1.11, 4.51) |
| Risk ratio: body immersion-no contact | 1.41 (1.07, 2.26) | N/a | 1.58 (1.12, 2.79) | 1.34 (0.87, 2.63) | 1.36 (1.00, 2.28) | 1.51 (1.08, 2.76) | 1.32 (1.04, 2.06) |
| Risk ratio: minimal contact-no contact | 1.10 (1.00, 1.62) | N/a | 1.10 (1.01, 1.61) | 1.08 (0.97, 1.72) | 1.09 (1.00, 1.60) | 1.22 (1.01, 2.14) | 1.08 (1.00, 1.49) |
| FIB overall effect[b] | 0.007 (-0.003, 0.025) | N/a | 0.003 (-0.003, 0.014) | 0.004 (-0.002, 0.022) | 0.005 (-0.005, 0.025) | 0.006 (-0.003, 0.025) | 0.009 (-0.008, 0.037) |

| | | | | | | | |
|---|---|---|---|---|---|---|---|
| FIB:swallowed water interaction[b] | 0.013 (-0.006, 0.051) | N/a | 0.007 (-0.005, 0.036) | 0.007 (-0.004, 0.040) | 0.008 (-0.011, 0.044) | 0.012 (-0.005, 0.051) | 0.017 (-0.014, 0.072) |
| FIB:body immersion interaction[b] | 0.005 (-0.004, 0.020) | N/a | 0.001 (-0.004, 0.009) | 0.003 (-0.002, 0.019) | 0.004 (-0.005, 0.021) | 0.004 (-0.004, 0.018) | 0.006 (-0.009, 0.030) |
| FIB:minimal contact interaction[b] | 0.002 (-0.007, 0.013) | N/a | 0.000 (-0.006, 0.004) | 0.002 (-0.003, 0.014) | 0.002 (-0.007, 0.015) | 0.002 (-0.006, 0.012) | 0.002 (-0.015, 0.021) |
| FIB effect at median time in water (min)[b] | N/a | 0.003 (-0.003, 0.014) | N/a | N/a | N/a | N/a | N/a |
| FIB effect at 95th percentile time in water (min) [b] | N/a | 0.001 (-0.014, 0.022) | N/a | N/a | N/a | N/a | N/a |
| Time in the water (min) [b] | N/a | 0.002 (-0.001, 0.009) | N/a | N/a | N/a | N/a | N/a |

[a] Weaker priors included Normal(0, 5) instead of Normal(0, 1.5) for beta parameters and Dirichlet(1,1,1) instead of Dirichlet(1,2,3) for the monotonic water contact effect.

[b] Effects are on the log transformed, mean-centered and standardized scale and contrast across the interquartile range (IQR) of FIB values

Figure S1: Directed acyclic graph for the water contact association with AGI to guide the model-building process in this study.

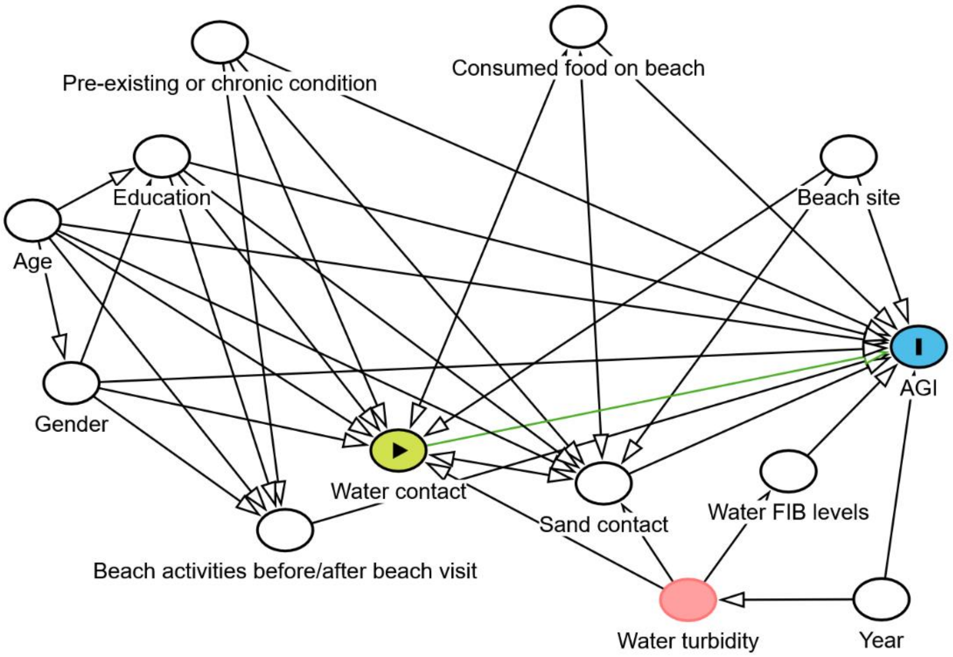

Figure S2: Summary plots of parameter posterior distributions and MCMC sampling chains from Model 1b.

**Intercept**

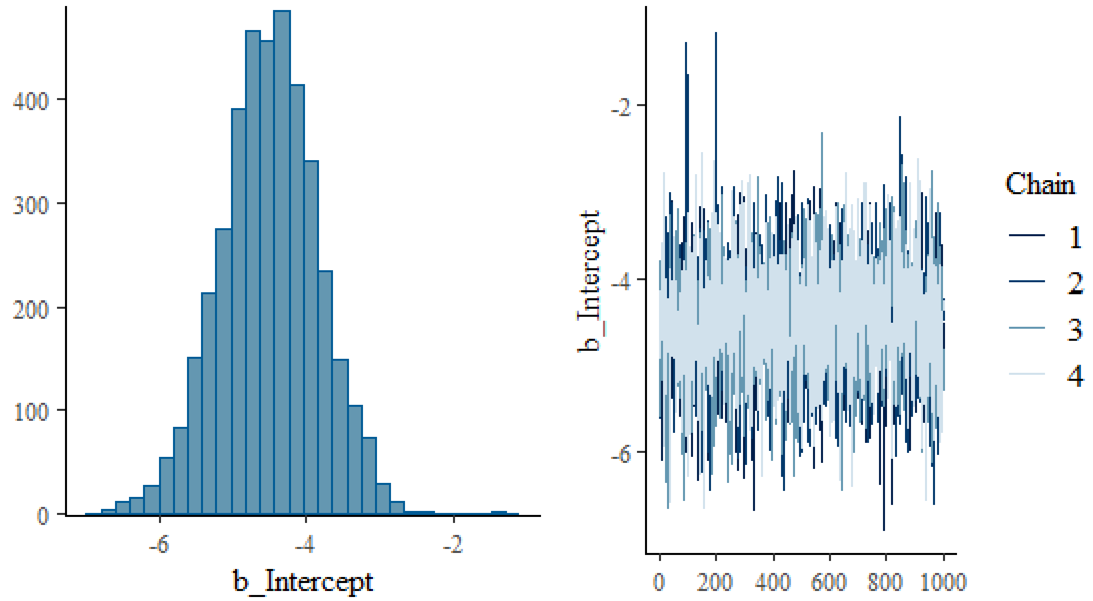


**Log *E. coli* maximum single sample value (beta parameter)**

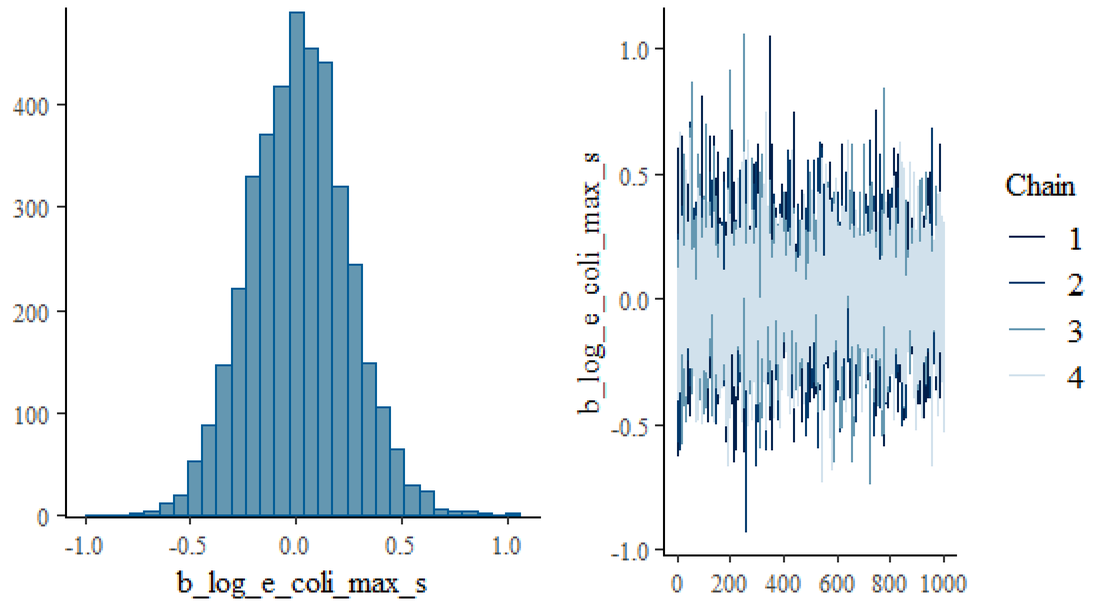

**Age: 5-9 years group (beta parameter)**

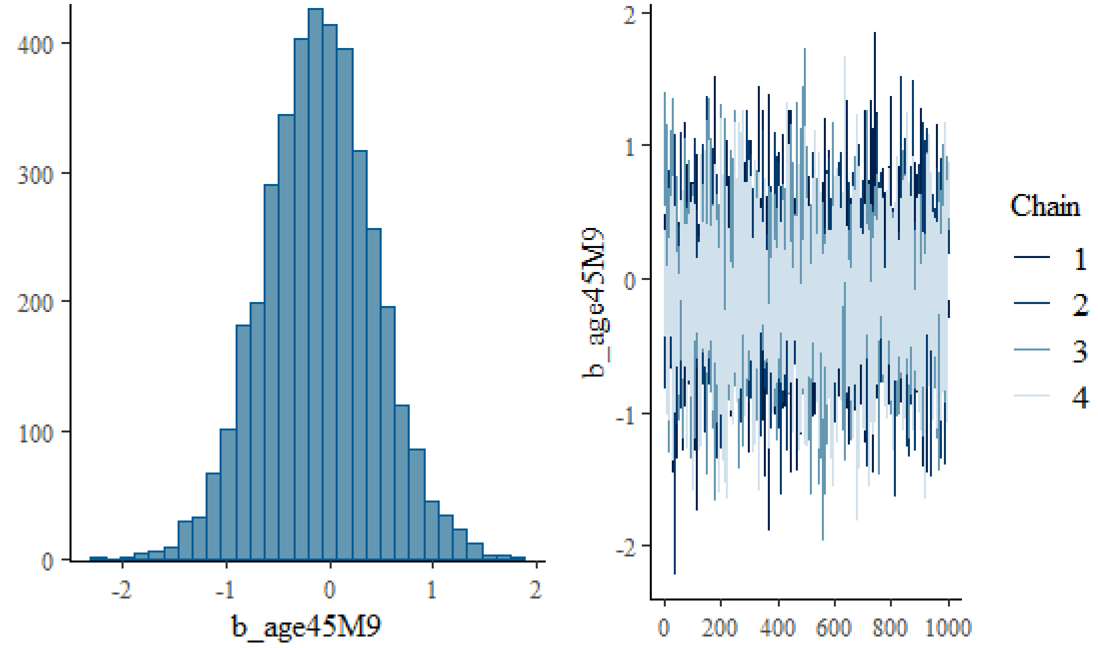


**Age: 10-14 years group (beta parameter)**

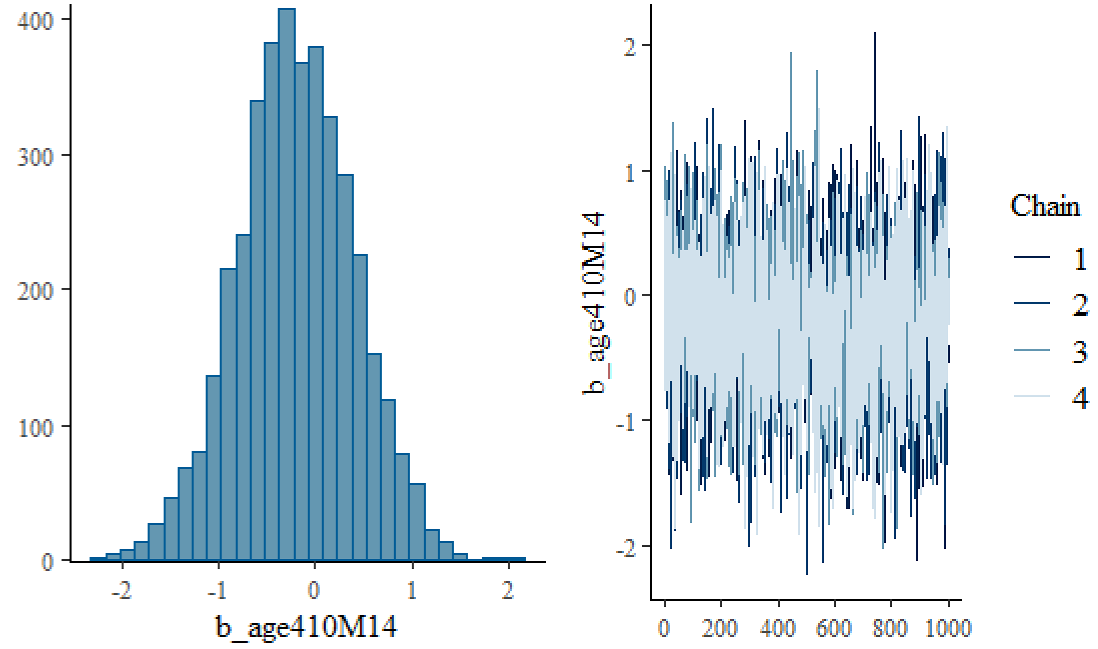

**Age: 15-19 years group (beta parameter)**

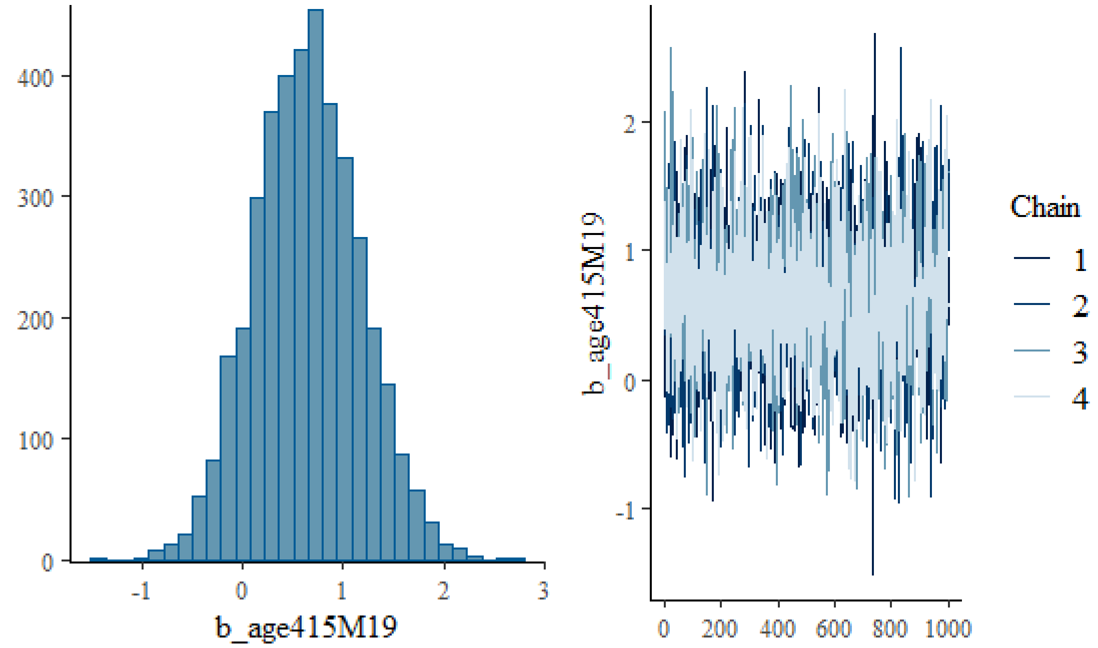


**Age: 20+ years group (beta parameter)**

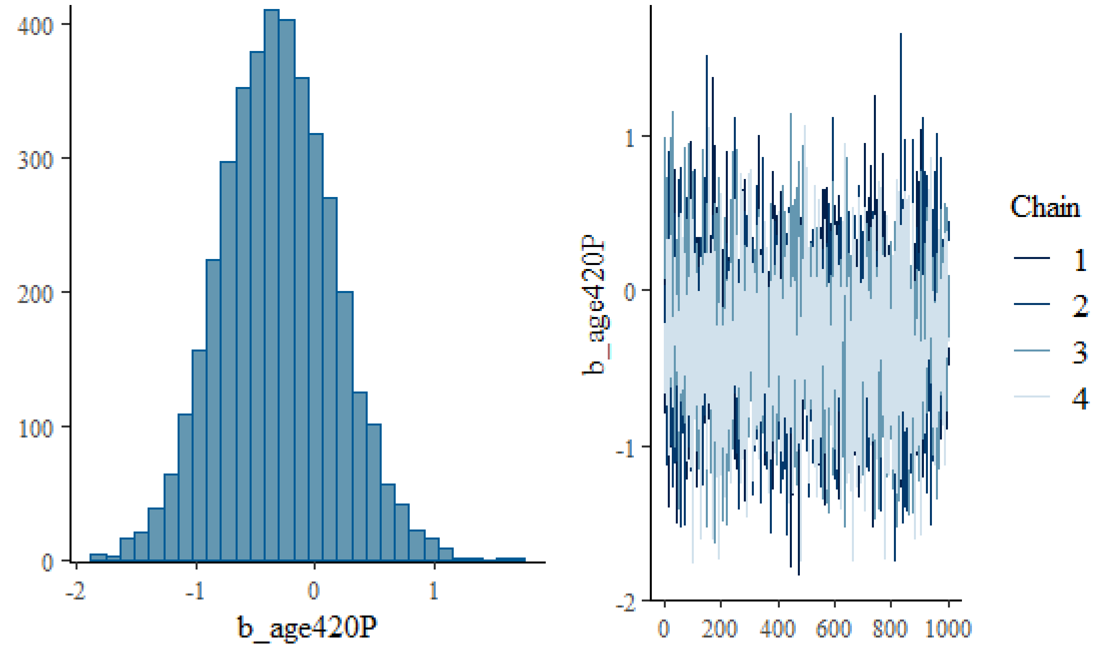

**Gender: Woman/girl group (beta parameter)**

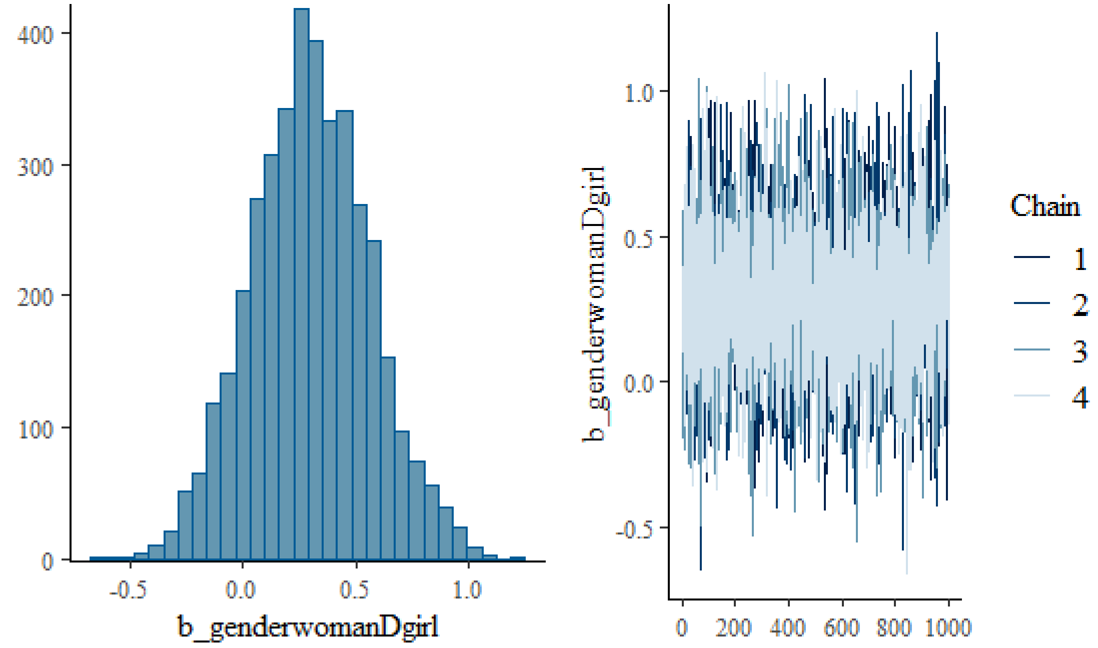


**Gender: Fluid/trans group (beta parameter)**

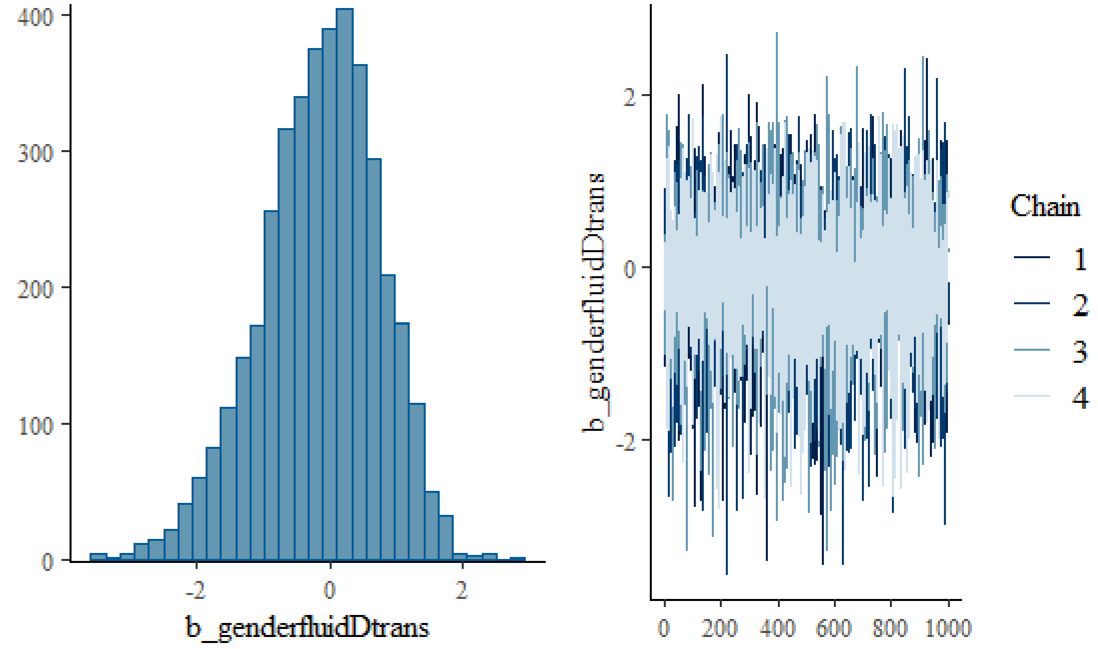

**Education: College/trades degree group (beta parameter)**

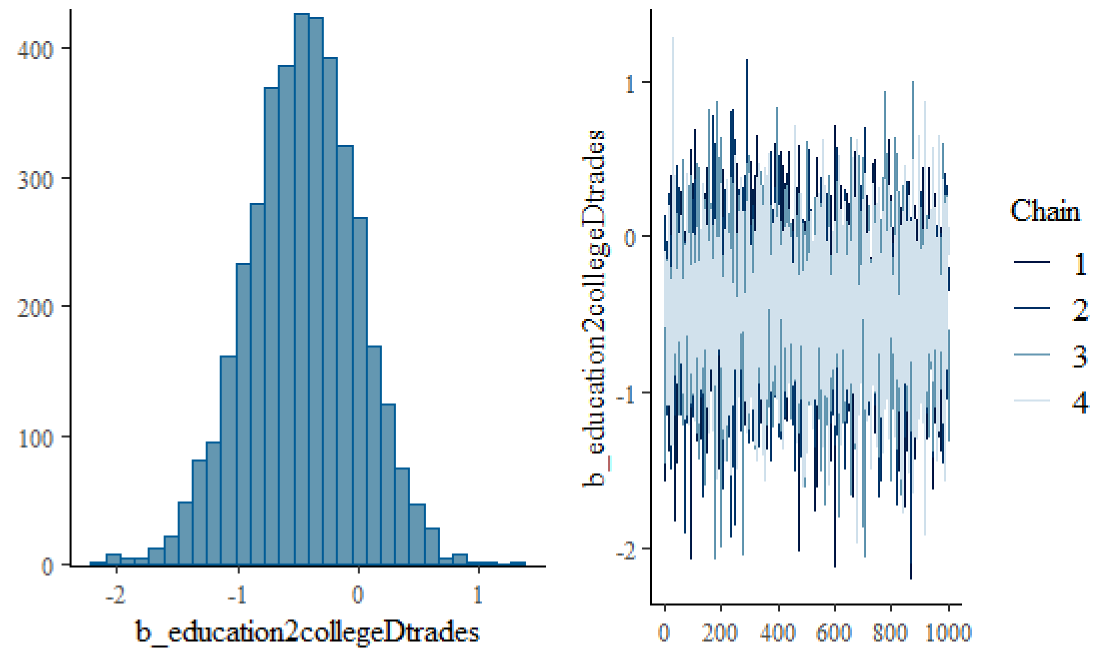


**Education: Bachelor's degree group (beta parameter)**

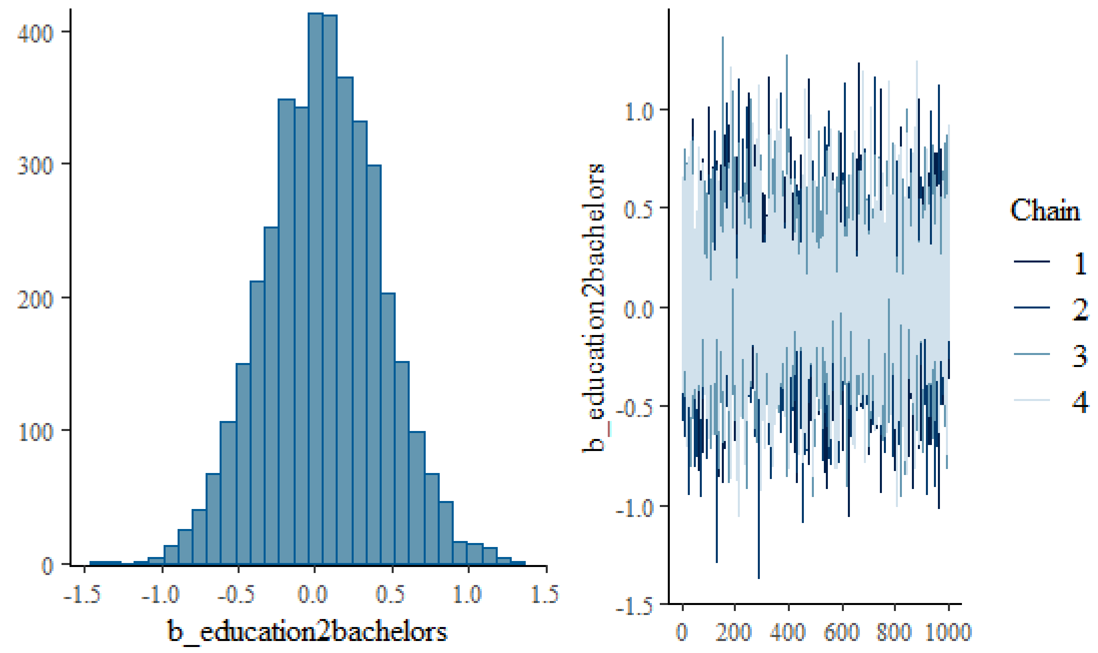

**Education: Post-graduate degree group (beta parameter)**

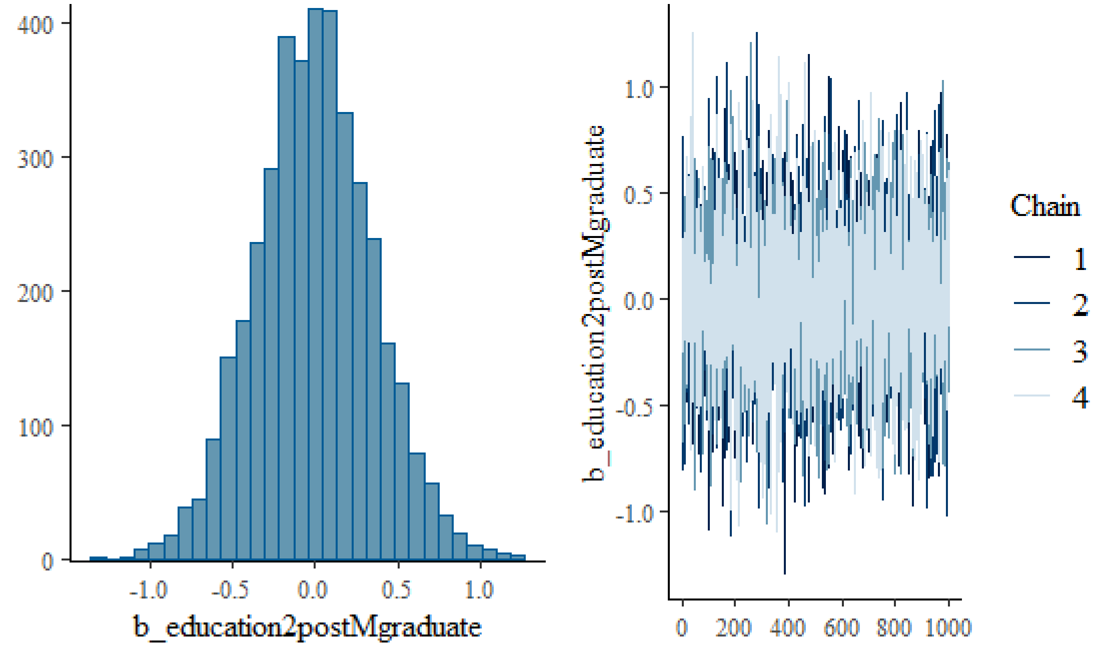


**GI condition: yes (beta parameter)**

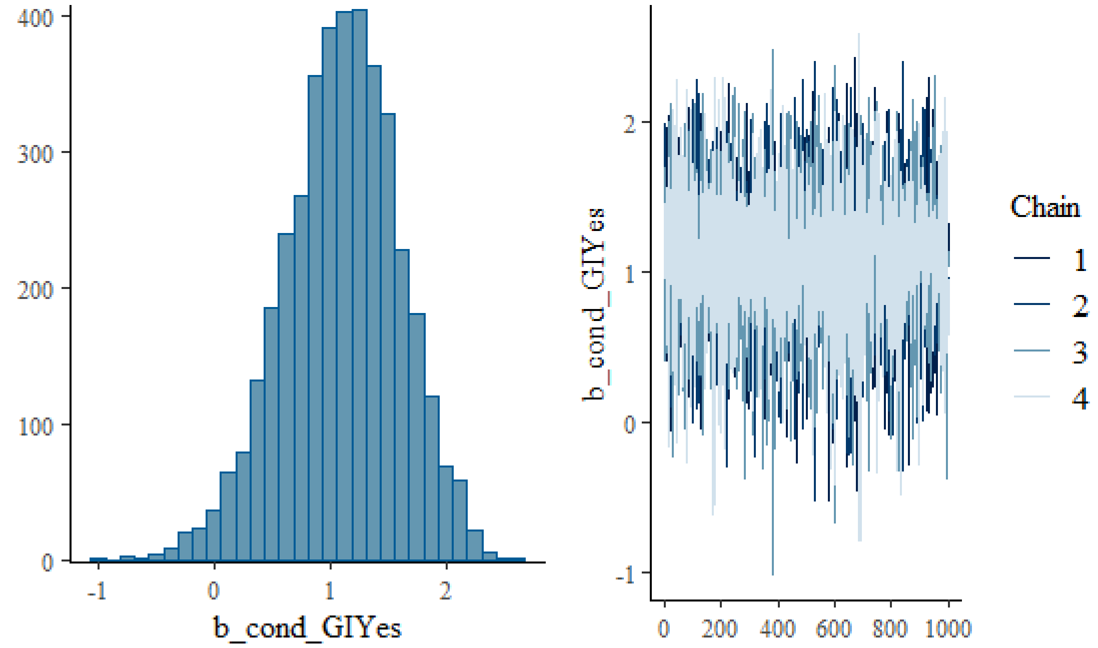

**Immune-condition: yes (beta parameter)**

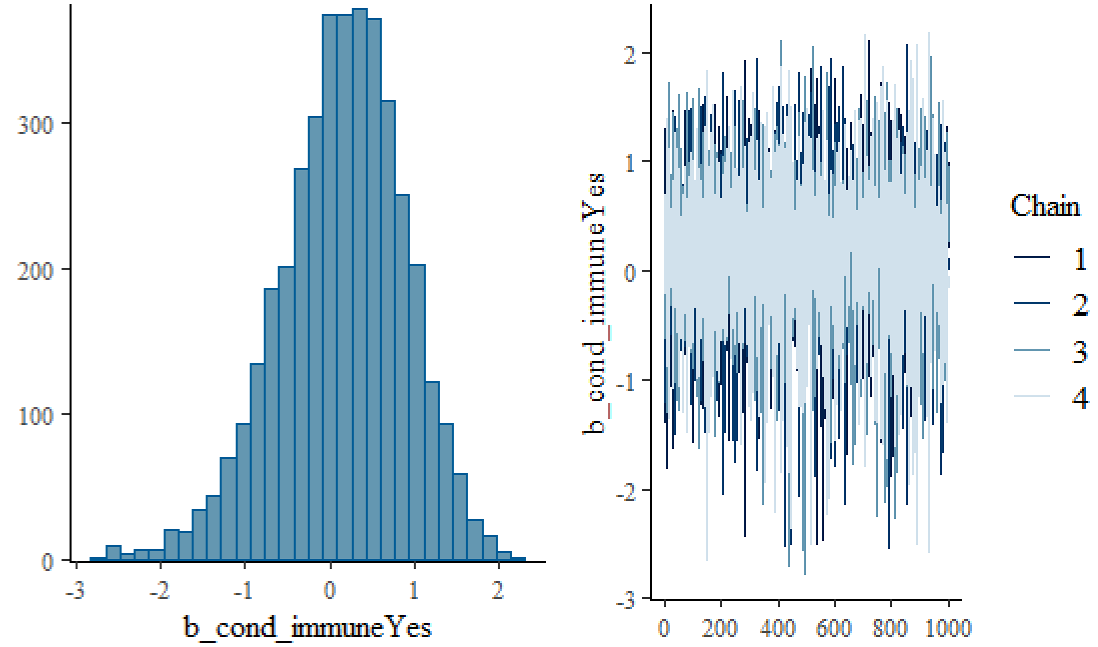


**Allergies: yes (beta parameter)**

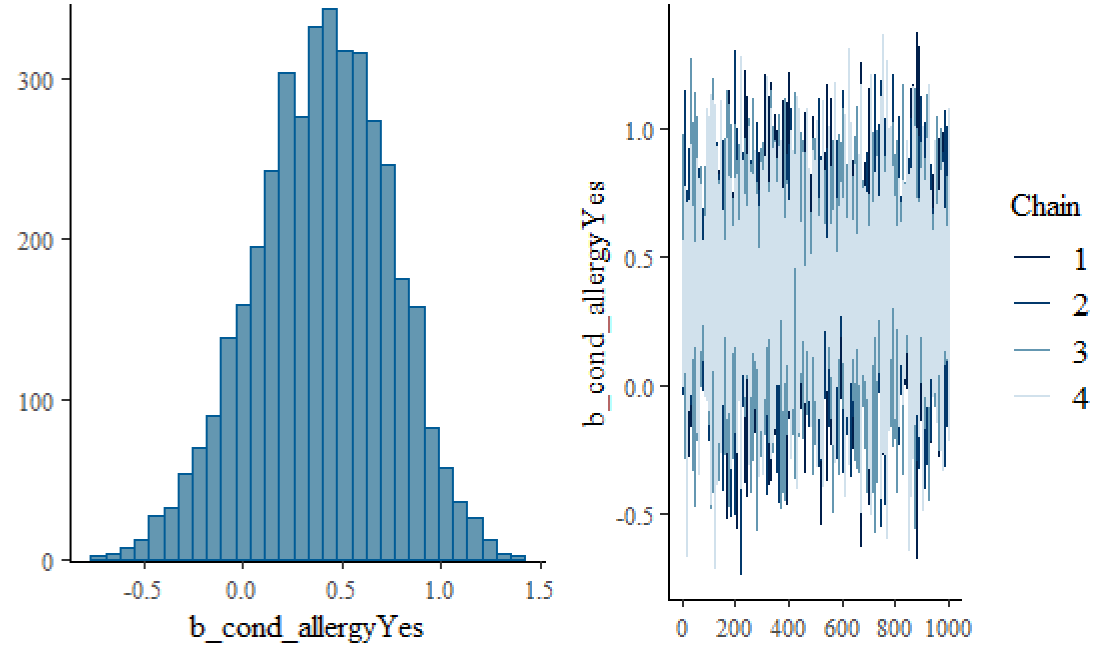

**Engaged in other recreational water activities: yes (beta parameter)**

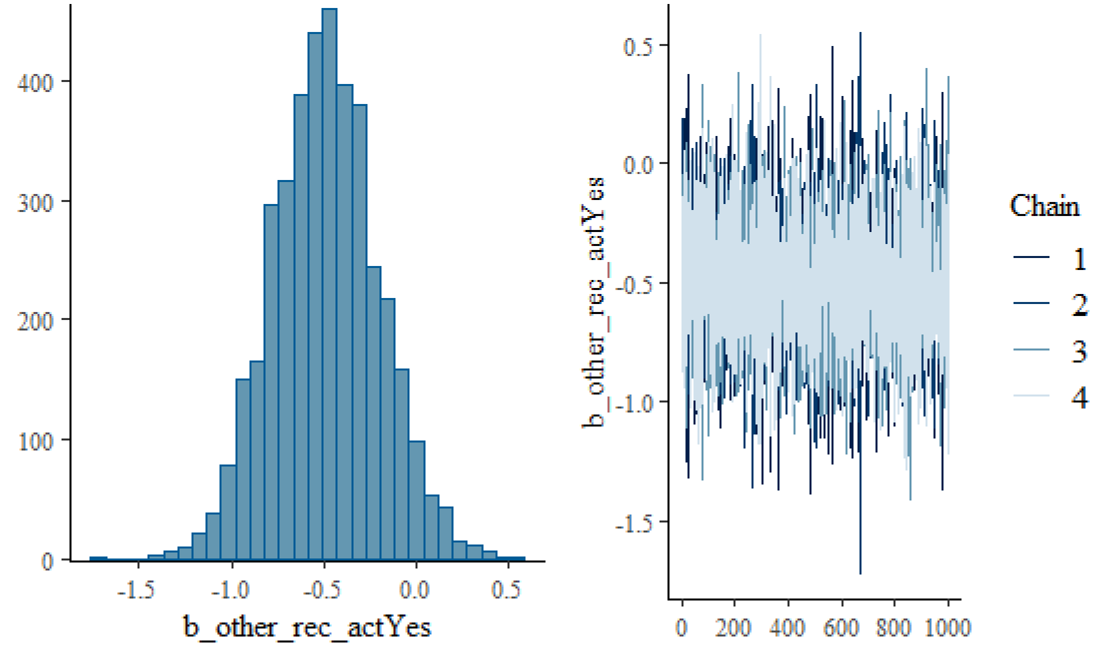


**Consumed food on the beach: yes (beta parameter)**

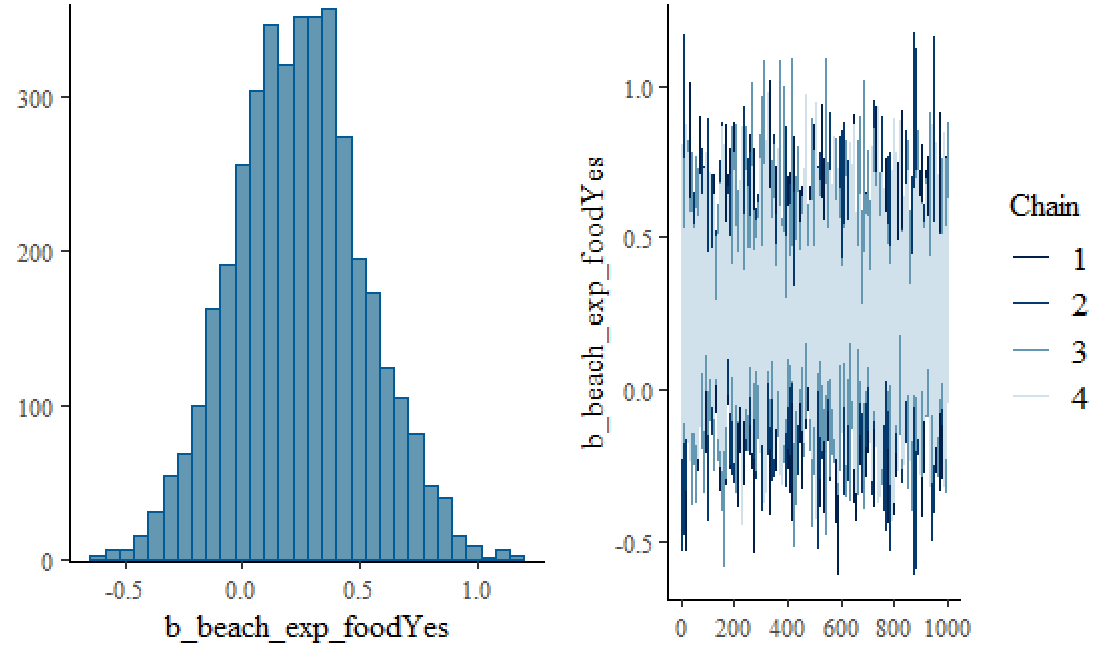

**Sand contact at the beach: yes (beta parameter)**

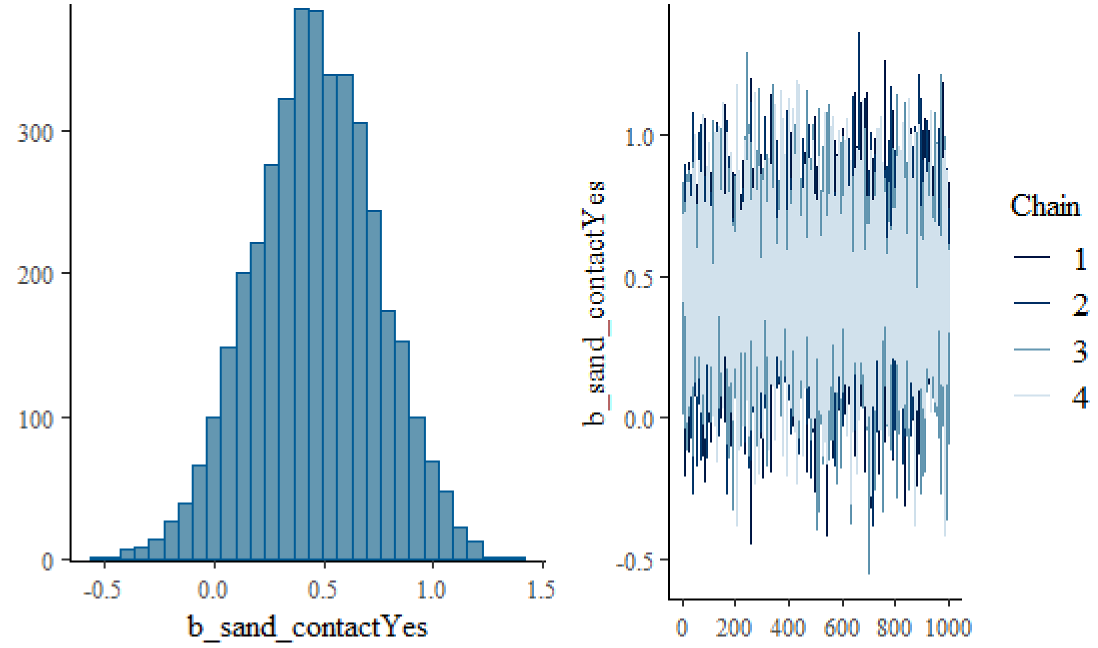


**Part of household group of participants: yes (beta parameter)**

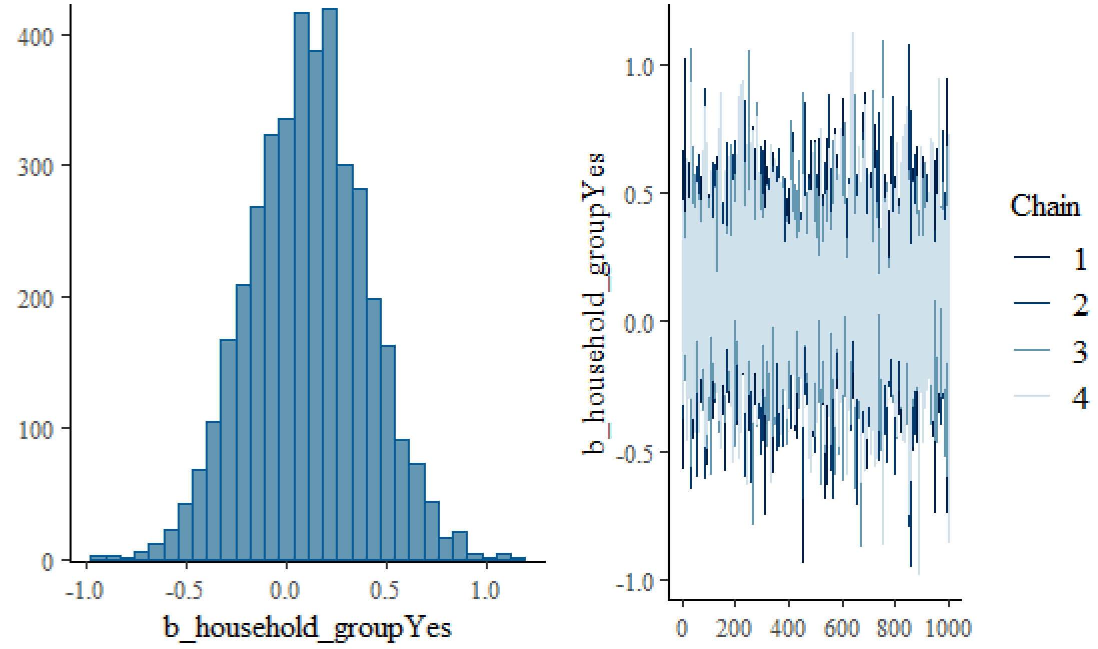

**Water contact average effect (beta parameter)**

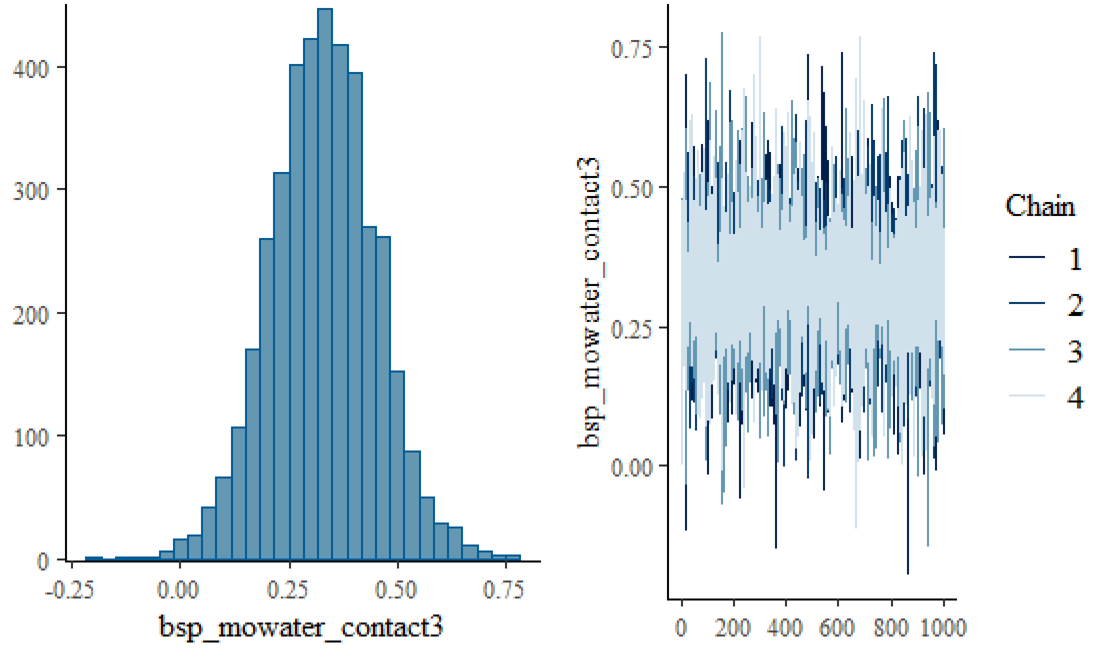


**Water contact-log *E. coli* interaction term (beta parameter)**

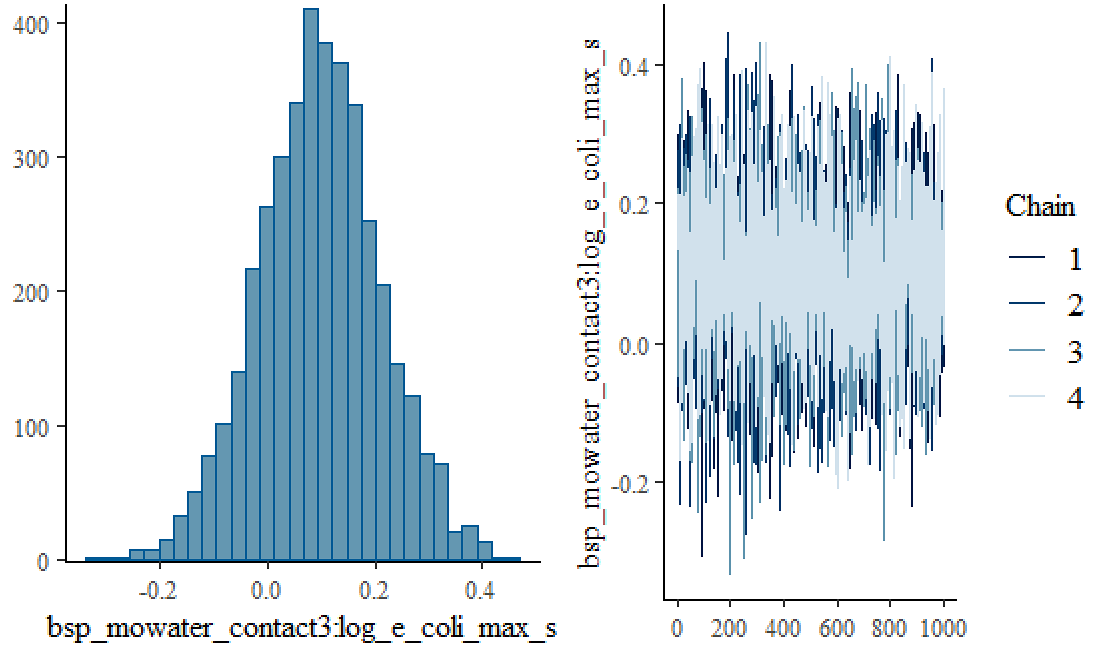

**Site random intercept (SD parameter)**

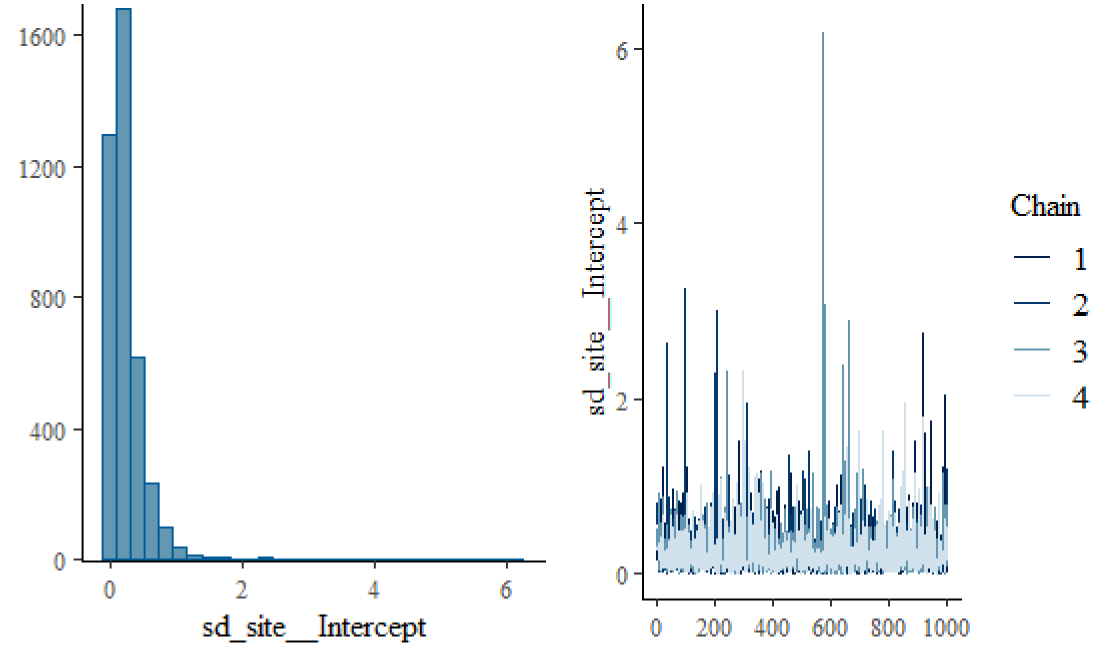


**Beach (within site) random intercept (SD parameter)**

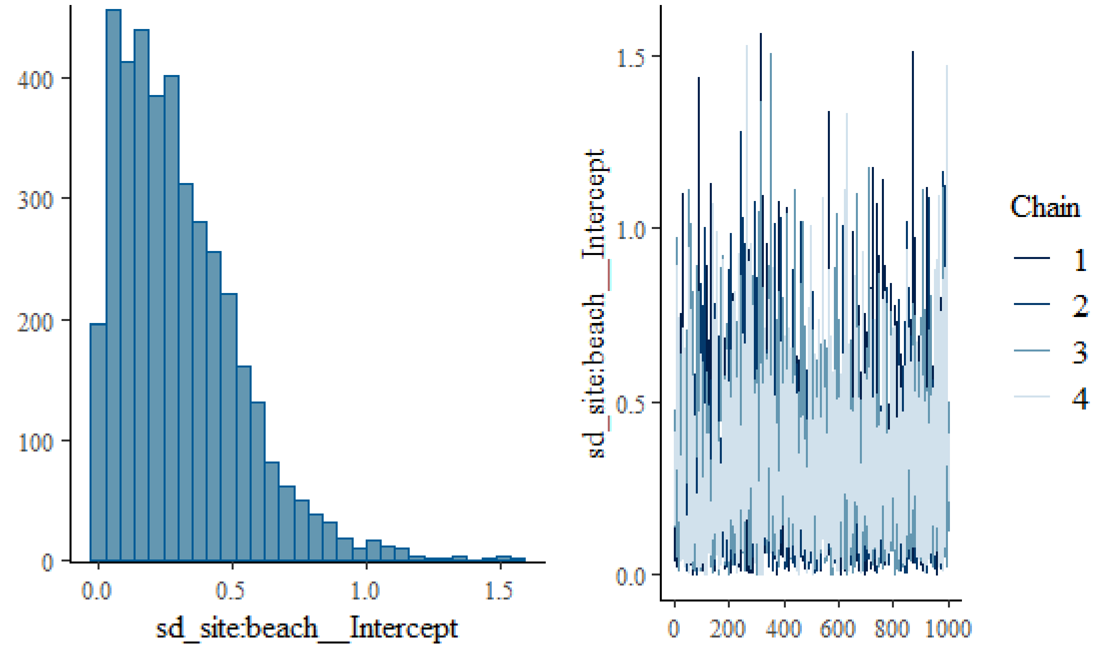

**Location-recruitment date (within beach within site) random intercept (SD parameter)**

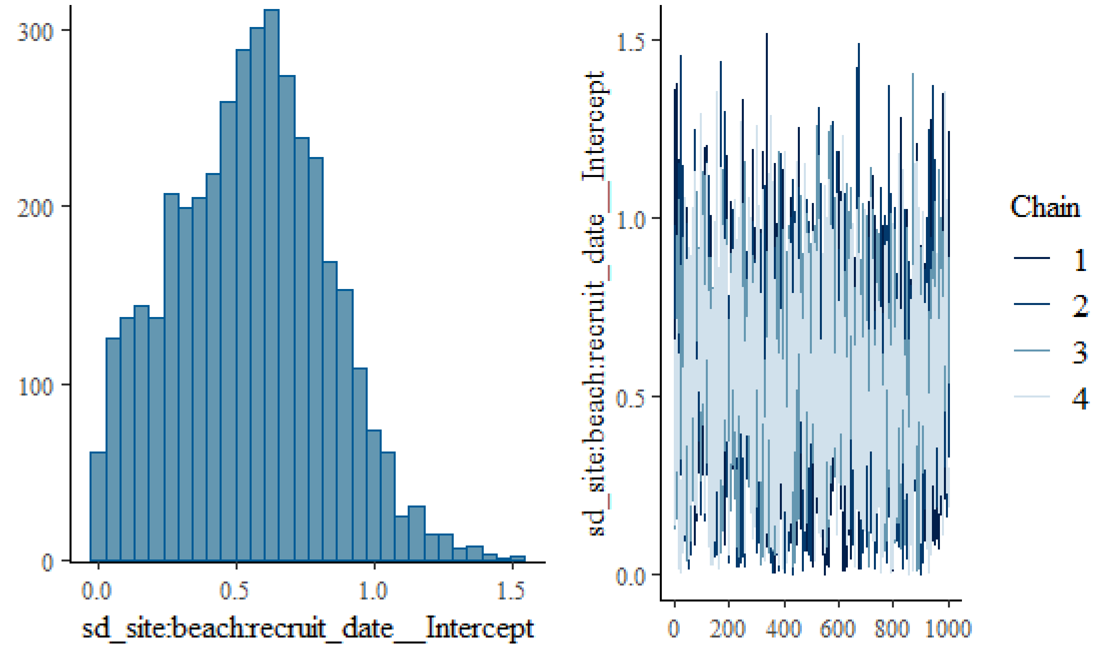

Figure S3: PAV-adjusted calibration plot for Model 1b.

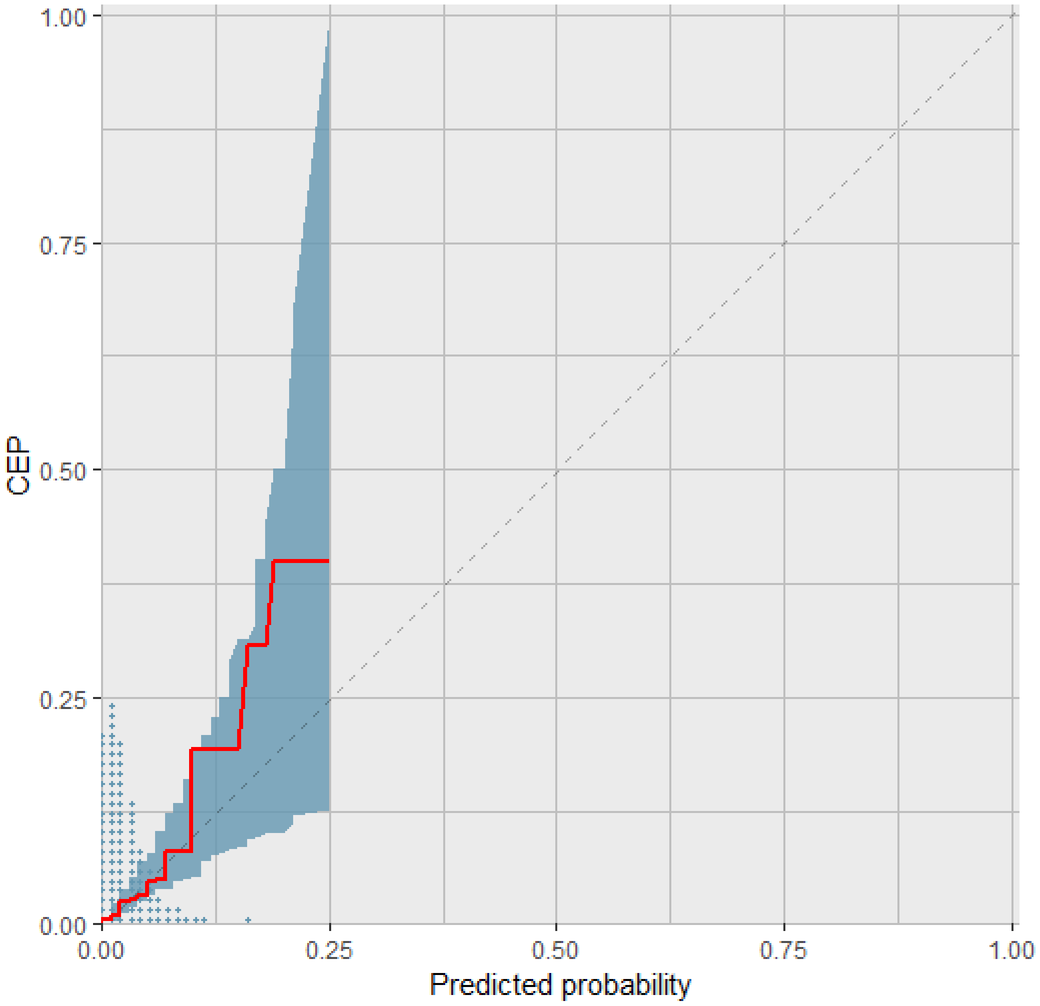

Figure S4: Summary of the predicted increase in incident risk of AGI per 1000 beachgoers by level of water contact (vs. no contact) stratified by site.

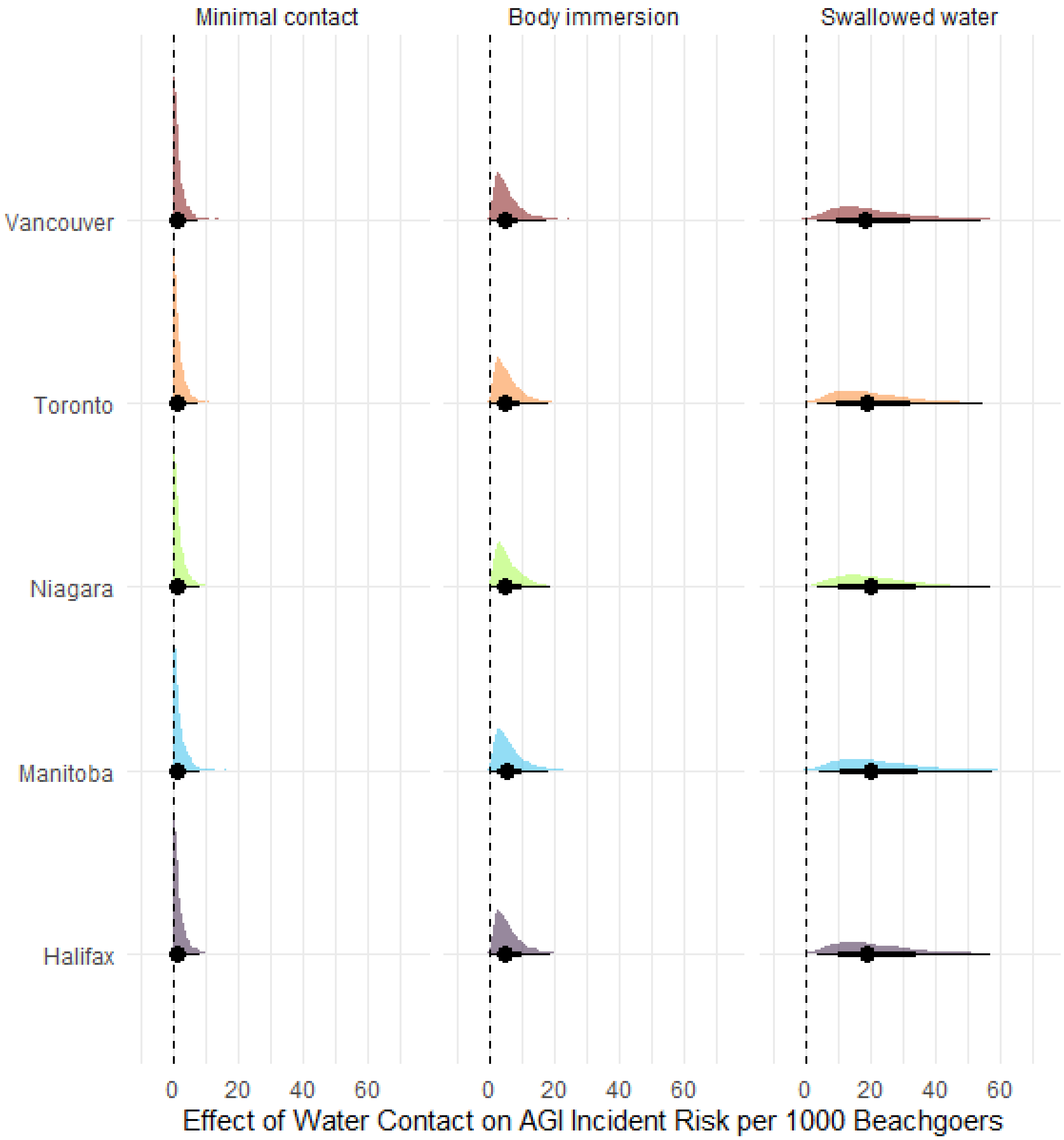

Figure S5: Effects of *E. coli* levels on the predicted probability of AGI by level of water contact for sensitivity analysis models. A) Time in water as alternative exposure; B) Diarrhea as alternative outcome; C) 3-day alternative follow-up period; D) 5-day alternative follow-up period; E) Weaker (less informative) priors; F) One randomly selected person per household.

**A)**

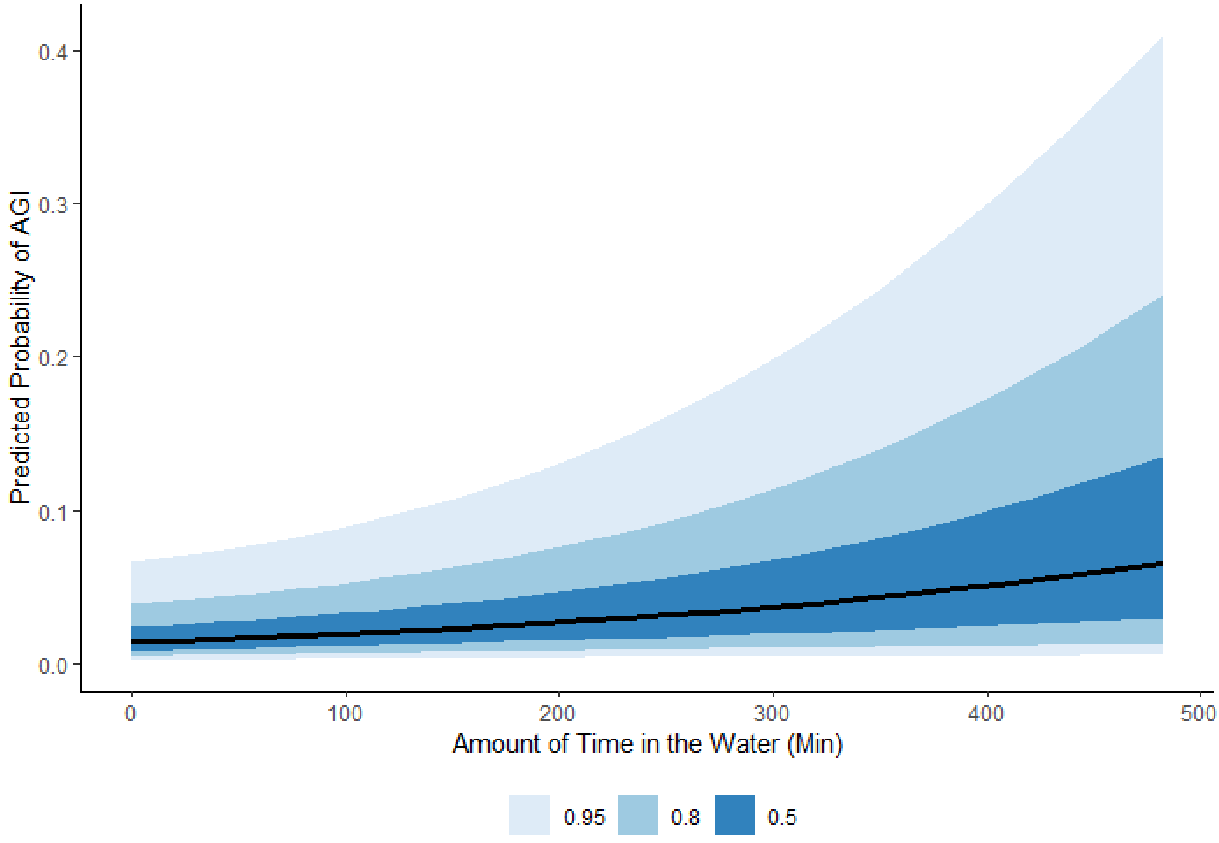

**B)**

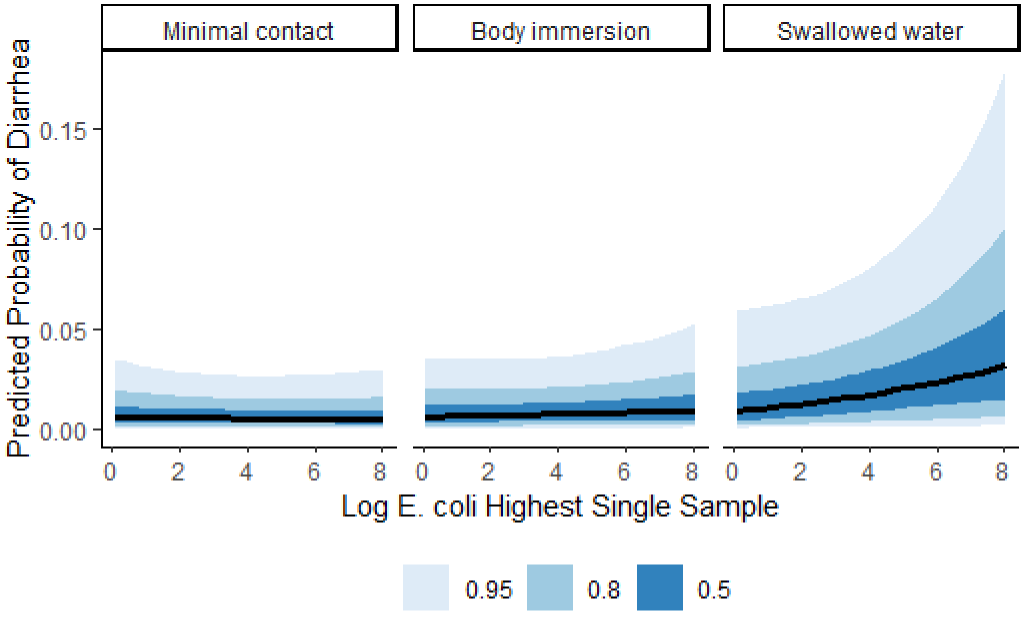

Minimal contact
Body immersion
Swallowed water
Predicted Probability of Diarrhea
0.00
0.05
0.10
0.15
0
2
4
6
8
Log E. coli Highest Single Sample
0.95
0.8
0.5


**C)**

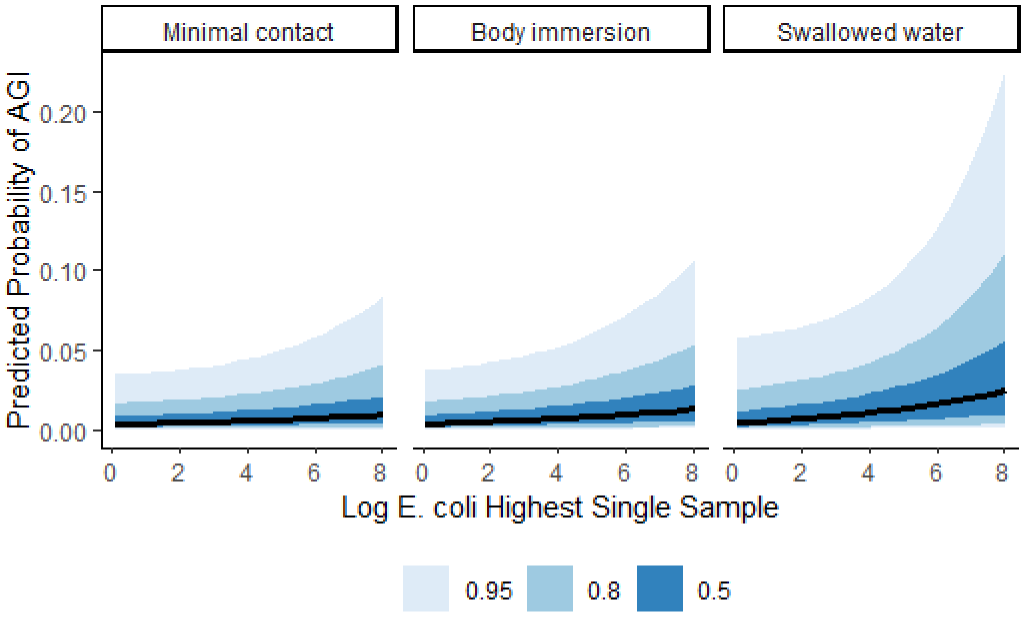

Minimal contact
Body immersion
Swallowed water
Predicted Probability of AGI
0.00
0.05
0.10
0.15
0.20
0
2
4
6
8
Log E. coli Highest Single Sample
0.95
0.8
0.5

D)

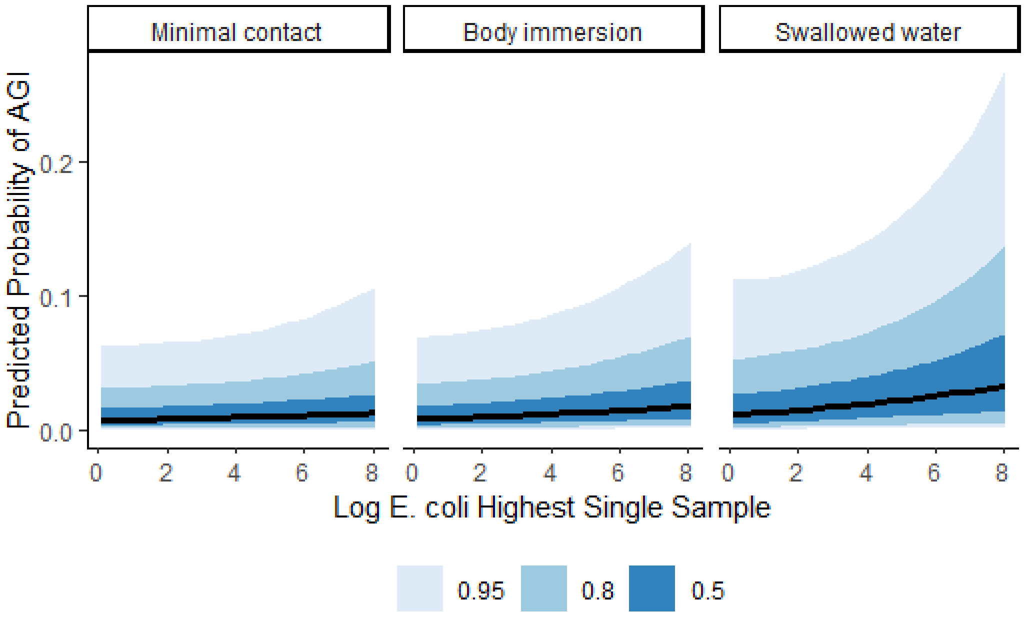
Minimal contact
Body immersion
Swallowed water
Predicted Probability of AGI
0.0
0.1
0.2
0
2
4
6
8
Log E. coli Highest Single Sample
0.95
0.8
0.5

E)

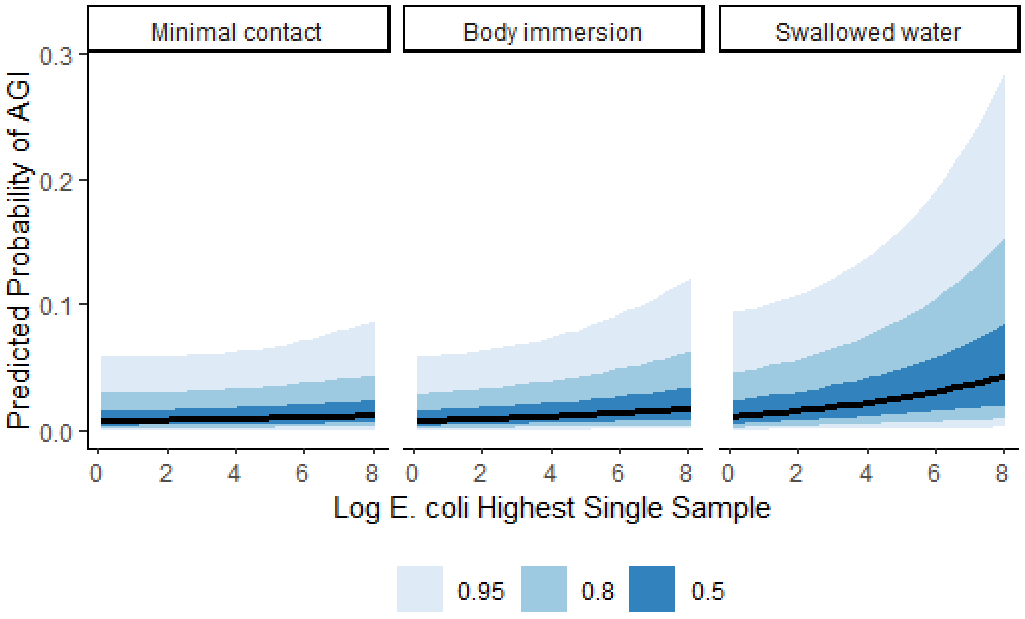
Minimal contact
Body immersion
Swallowed water
Predicted Probability of AGI
0.0
0.1
0.2
0.3
0
2
4
6
8
Log E. coli Highest Single Sample
0.95
0.8
0.5

**F)**

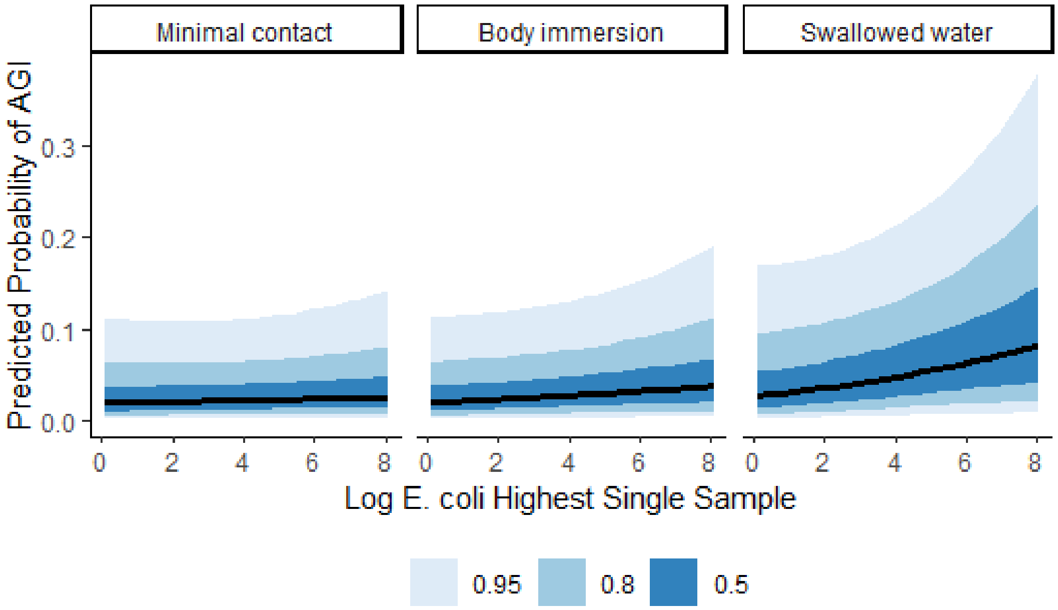

Minimal contact
Body immersion
Swallowed water
Predicted Probability of AGI
0.0
0.1
0.2
0.3
0
2
4
6
8
Log E. coli Highest Single Sample
0.95
0.8
0.5

Figure S6: Summary of the predictive effect of *E. coli* on AGI incident risk in negative control analysis model that contained only participants that did not report any water contact.

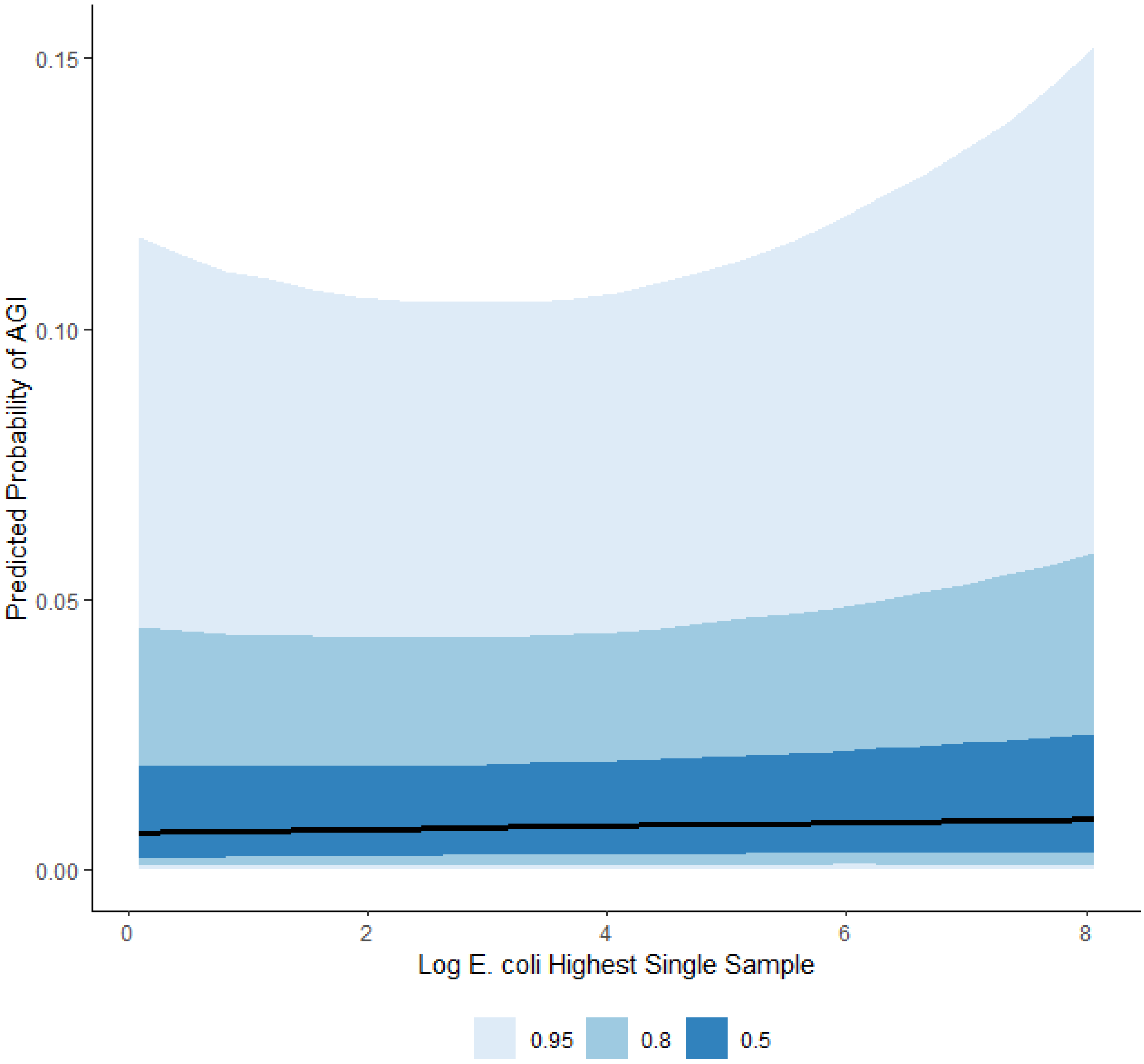
